# Diabetes following SARS-CoV-2 infection: Incidence, persistence, and implications of COVID-19 vaccination. A cohort study of fifteen million people

**DOI:** 10.1101/2023.08.07.23293778

**Authors:** Kurt Taylor, Sophie Eastwood, Venexia Walker, Genevieve Cezard, Rochelle Knight, Marwa Al Arab, Yinghui Wei, Elsie M F Horne, Lucy Teece, Harriet Forbes, Alex Walker, Louis Fisher, Jon Massey, Lisa E M Hopcroft, Tom Palmer, Jose Cuitun Coronado, Samantha Ip, Simon Davy, Iain Dillingham, Caroline Morton, Felix Greaves, John Macleod, Ben Goldacre, Angela Wood, Nishi Chaturvedi, Jonathan A C Sterne, Rachel Denholm, The Longitudinal Health and Wellbeing and Data and Connectivity UK COVID-19 National Core Studies, CONVALESCENCE study, The OpenSAFELY collaborative

## Abstract

**Background:** Type 2 diabetes (T2DM) incidence is increased after diagnosis of COVID-19. The impact of vaccination on this increase, for how long it persists, and the effect of COVID-19 on other types of diabetes remain unclear.

**Methods:** With NHS England approval, we studied diabetes incidence following COVID-19 diagnosis in pre-vaccination (N=15,211,471, January 2020-December 2021), vaccinated (N =11,822,640), and unvaccinated (N=2,851,183) cohorts (June-December 2021), using linked electronic health records. We estimated adjusted hazard ratios (aHRs) comparing diabetes incidence post-COVID-19 diagnosis with incidence before or without diagnosis up to 102 weeks post-diagnosis. Results were stratified by COVID-19 severity (hospitalised/non-hospitalised) and diabetes type.

**Findings:** In the pre-vaccination cohort, aHRS for T2DM incidence after COVID-19 (compared to before or without diagnosis) declined from 3.01 (95% CI: 2.76,3.28) in weeks 1-4 to 1.24 (1.12,1.38) in weeks 53-102. aHRS were higher in unvaccinated than vaccinated people (4.86 (3.69,6.41)) versus 1.42 (1.24,1.62) in weeks 1-4) and for hospitalised COVID-19 (pre-vaccination cohort 21.1 (18.8,23.7) in weeks 1-4 declining to 2.04 (1.65,2.51) in weeks 52-102), than non-hospitalised COVID-19 (1.45 (1.27,1.64) in weeks 1-4, 1.10 (0.98,1.23) in weeks 52-102). T2DM persisted for 4 months after COVID-19 for ∼73% of those diagnosed. Patterns were similar for Type 1 diabetes, though excess incidence did not persist beyond a year post-COVID-19.

**Interpretation:** Elevated T2DM incidence after COVID-19 is greater, and persists longer, in hospitalised than non-hospitalised people. It is markedly less apparent post-vaccination. Testing for T2DM after severe COVID-19 and promotion of vaccination are important tools in addressing this public health problem.

**Research in context:** *Evidence before this study:* We searched PubMed for population-based observational studies published between December 1st 2019 and July 12th 2023 examining associations between SARS-CoV-2 infection or COVID-19 diagnosis (search string: SARS-CoV-2 or COVID* or coronavirus*) and subsequent incident diabetes (search term: diabetes). Of nineteen relevant studies; eight had a composite outcome of diabetes types, six stratified by diabetes type and five pertained to type-1-diabetes (T1DM) only. We did not identify any studies relating to gestational or other types of diabetes. Eleven studies were from the US, three from the UK, two from Germany, one from Canada, one from Denmark and one from South Korea. Most studies described cumulative relative risks (for infection versus no infection) one to two years post-SARS-CoV-2 infection of 1.2 to 2.6, though four studies found no associations with T1DM after the post-acute period. All studies lacked the power to compare diabetes relative risk by type, severity, and vaccination status in population subgroups. One study examined relative risks by vaccination status, but this used a composite outcome of diabetes and hyperlipidaemia and was conducted in a predominantly white male population. Two studies of T1DM found no evidence of elevated risk beyond 30 days after COVID-19 diagnosis, whilst two reported elevated risks at six months. Two studies of type 2 diabetes (T2DM) examined relative risks by time period post-infection: one study of US insurance claims reported a persistent association six months post-infection, whereas a large UK population-based study reported no associations after 12 weeks. However, the latter study used only primary care data, therefore COVID-19 cases were likely to have been under-ascertained. No large studies have investigated the persistence of diabetes diagnosed following COVID-19; key to elucidating the role of stress/steroid-induced hyperglycaemia.

*Added value of this study:* This study, which is the largest to address the question to date, analysed linked primary and secondary care health records with SARS-CoV-2 testing and COVID-19 vaccination data for 15 million people living in England. This enabled us to compare the elevation in diabetes incidence after COVID-19 diagnosis by diabetes type, COVID-19 severity and vaccination status, overall and in population subgroups. Importantly, excess diabetes incidence by time period since infection could also be quantified. Since healthcare in the UK is universal and free-at-the-point-of-delivery, almost the entire population is registered with primary care. Therefore the findings are likely to be generalisable. We found that, before availability of COVID-19 vaccination, a COVID-19 diagnosis (vs. no diagnosis) was associated with increased T2DM incidence which remained elevated by approximately 30% beyond one year after diagnosis. Though still present (with around 30% excess incidence at eight weeks), these associations were substantially attenuated in unvaccinated compared with vaccinated people. Excess incidence was greater in people hospitalised with COVID-19 than those who were not hospitalised after diagnosis. T1DM incidence was elevated up to, but not beyond, a year post COVID-19. Around 73% of people diagnosed with incident T2DM after COVID-19 still had evidence of diabetes four months after infection.

*Implications of all the available evidence:* There is a 30-50% elevated T2DM incidence post-COVID-19, but we report the novel finding that there is elevated incidence beyond one-year post-diagnosis. Elevated T1DM incidence did not appear to persist beyond a year, which may explain why previous studies disagree. For the first time in a general-population dataset, we demonstrate that COVID-19 vaccination reduces, but does not entirely ameliorate, excess diabetes incidence after COVID-19. This supports a policy of universal vaccination and suggests that other public health activities, such as enhanced diabetes screening after severe COVID-19, may be warranted, particularly in unvaccinated people.

## Introduction

At least 660 million people worldwide have been infected with severe acute respiratory syndrome coronavirus 2 (SARS-CoV-2).^1, 2^ Therefore reports of excess diabetes risk after COVID-19^3–13^ have alarming public health implications. A 30% to 50% excess of type-2-diabetes (T2DM) incidence following SARS-CoV-2 infection has been reported^3, 4, 6, 8, 13, 14^. In contrast, the only three studies sufficiently powered to examine type-1-diabetes (T1DM) reported no association with SARS-CoV-2 ^15–17^. A key question is whether excess diabetes after COVID-19 is driven by short-term causes of hyperglycaemia (stress/steroid-induced) or is a durable consequence of infection. Yet most studies only examined diabetes incidence at a fixed timepoint after COVID-19 ^3, 4, 9–12^ and those that stratified by time period post-infection do not concur ^3, 4, 6, 9, 11, 13^. COVID-19 severity is likely to be a key determinant of downstream sequelae ^4–11, 13, 15, 16^, only one study, in a predominantly white male cohort, examined the impact of vaccination on diabetes after COVID ^13^.

In the largest study to date, we quantified associations of COVID-19 with incident diabetes using a UK database of linked COVID-19 testing data and primary and secondary care records. We investigated how associations varied: at different timepoints up to two years post-diagnosis; by diabetes type; according to vaccination availability and vaccination status; by COVID-19 severity; and within population subgroups.

## Methods

Information on data access can be found under “Data Sharing” at the end of this paper. Analyses were performed according to a pre-specified protocol which can be found along with publicly available code lists and analysis code: https://github.com/opensafely/post-covid-diabetes. This study was approved by the Health Research Authority [REC reference 22/PR/0095] and by the University of Bristol’s Faculty of Health Sciences Ethics Committee [reference 117269] (see “Information governance and ethical approval”).

### Study design and data source

This observational cohort study included all adults registered with a primary care general practice using TPP software in England. Data were linked, stored and analysed securely within the OpenSAFELY platform (https://opensafely.org/), which includes individual-level, primary care electronic health records from 24 million people, covering around 40% of the English population, securely linked to the Second Generation Surveillance System (SGSS) for Pillar 1 and Pillar 2 SARS-COV-2 infection laboratory testing data, NHS hospital admissions (Secondary Uses Services data) and the Office of National Statistics death registry, including causes of death^18^. COVID-19 vaccination records (National Immunisation Management System) are available within TPP primary care data.

The UK’s COVID-19 vaccine roll-out started on December 8th, 2020 with timing of eligibility in order of priority groups determined by the Joint Committee on Vaccination and Immunisation, based on age, clinical vulnerability and health and social care occupation ^19^. All adults in England were eligible to receive a first vaccination by June 18th 2021, and a second vaccination by August 2021^20^.

### Outcomes

Diabetes phenotypes were defined using primary care and hospital admission data (**Supplementary Table 1**). An updated, clinician-verified SNOMED-CT and ICD-10 diabetes diagnostic adjudication algorithm^21, 22^ was used to define incident T1DM and T2DM, gestational diabetes, other or non-specific diabetes, and diabetes unlikely (**Supplementary Figure 1**). In the pre-vaccination cohort, to differentiate T2DM from temporary steroid- or stress-induced hyperglycaemia, a secondary outcome of ‘persistent T2DM’ was defined as T2DM with continued treatment (≥2 prescriptions of glucose-lowering medication) or elevated HbA1c levels (>= 47.5 mmol)) 4 months after diagnosis.

### Exposures

The date of COVID-19 diagnosis was defined as the earliest of the date of a positive SARS-CoV-2 polymerase chain reaction or antigen test in the SGSS system, a confirmed COVID-19 diagnosis in primary or secondary care, or death with SARS-CoV-2 infection listed as primary or underlying cause. People hospitalised with a COVID-19 diagnosis in the primary position within 28 days of first COVID-19 diagnosis were defined as ‘hospitalised COVID-19’, otherwise as ‘non-hospitalised COVID-19’.

### Potential confounding variables

Primary and secondary care records up to the cohort start date (see below) were used to define age, sex, ethnicity, socioeconomic deprivation (area-based index of multiple deprivation quintiles), smoking status and region. We defined additional potential confounding variables based on previous disease diagnoses, comorbidities and medications (**Supplementary Table 2**).

### Study population

We defined three cohorts, summarised in **Supplementary Table 3**. In the ‘pre-vaccination’ cohort, follow-up started on January 1^st^ 2020 (baseline) and ended on the earliest of December 14th 2021 (when the Omicron variant became dominant in England)^23^, date that the outcome event was recorded, and date of death. Exposure was defined as a COVID-19 diagnosis between baseline and the earliest of eligibility for COVID-19 vaccination (based on age and clinical vulnerability) and date of first vaccination: this exposure period was before the Delta variant became dominant in England. The other two cohorts were followed during the period when the Delta variant was dominant in England: between June 1^st^ 2021 (baseline) and December 14^th^ 2021 (study end date). Follow-up in the ‘vaccinated’ cohort started at the later of baseline and two weeks after a second COVID-19 vaccination and ended at the earliest of the study end date, outcome event date, and date of death. The ‘unvaccinated’ cohort had not received a COVID-19 vaccine by 12 weeks after they became eligible for vaccination. Follow-up started at the later of baseline and 12 weeks after eligibility for vaccination and ended at the earliest of the study end date, outcome event date, date of death, and date of first vaccination.

People eligible for each cohort had been registered with an English GP using TPP software for at least six months before the cohort baseline, were alive and aged between 18 and 110 years at baseline, and had known sex, region and area deprivation. People with a history of SARS-CoV-2 infection or COVID-19 diagnosis before the cohort baseline were excluded. In the vaccinated cohort, people who received a COVID-19 vaccination before December 8^th^ 2020, or a second dose before or less than three weeks after their first dose, or received more than one type of vaccine before 7^th^ May 2021, were excluded. In the unvaccinated cohort, people who could not be assigned to a vaccination priority group as defined by the Joint Committee on Vaccination and Immunisation (JCVI) were excluded.

People were excluded if they had previously been diagnosed with diabetes prior to the study start date. For gestational diabetes, the study population was restricted to women.

### Statistical analyses

We described baseline demographic and clinical characteristics of each cohort and calculated the number of events per outcome, person-years of follow-up and incidence rates (per 100,000 person-years) of events before and after all, hospitalised and non-hospitalised COVID-19.

For each outcome, we analysed time to first event. Cox models were fitted with calendar time scale using the cohort-specific baseline as the origin (time zero). We estimated hazard ratios (HRs) comparing follow-up after COVID-19 with follow-up before or without COVID-19, splitting follow-up time into periods 1-4 weeks and 5-28 weeks after COVID-19 for all cohorts and additionally 29-52 and 53-102 weeks after COVID-19 for the pre-vaccination cohort. All models were stratified by region, to account for between-region variation. For each outcome and cohort, we estimated: (i) age and sex; and (ii) maximally (including all potential confounders) adjusted HRs. We conducted subgroup analyses according to whether people had been hospitalised for COVID-19 within 28 days of COVID-19 diagnosis. For T2DM only, we conducted additional subgroup analyses by age group (18-39 / 40-59 / 60-79 / 80-110 years), sex, ethnicity, prior history of pre-diabetes and obesity. Absolute excess risks (AER) of T2DM after COVID-19, weighted by the proportions of individuals in age and sex strata in the pre-vaccination cohort, were derived. Further details of the statistical analyses and AER calculations are provided in the supplementary material.

### Sensitivity analyses

We performed sensitivity analyses for T2DM outcomes only. We repeated the main analysis in people who had COVID-19 before the cohort start date (vaccinated and unvaccinated cohorts only), and separating events on day 0 (day of COVID-19 diagnosis) from the rest of weeks 1-4, to explore the influence of simultaneous reporting of the exposure and outcome.

## Results

**Supplementary Figures 2-4** show participant selection into each cohort. Among 15,211,471 people in the pre-vaccination cohort, 827,074 were diagnosed with COVID-19 during the study period (**Table 1**). Corresponding numbers were 750,370 diagnosed with COVID-19 among 11,822,640 people in the vaccinated cohort and 147,044 among 2,851,183 in the unvaccinated cohort. Compared to the vaccinated cohort, people who remained unvaccinated were younger (61.5% versus 28.3% aged ≤40 years in the unvaccinated and vaccinated cohorts, respectively), more likely to be men (59% versus 47.3% respectively), and more likely to be from South Asian (9.6% versus 5.1%, respectively), Black (5.1% versus 1.5%, respectively) and other (5.6% and 1.7%) ethnic backgrounds, and the most deprived background (29.9% versus 15.7%, respectively). People in the unvaccinated cohort had fewer comorbidities compared to those in the vaccinated cohort (**Supplementary Table 4**).

**Table 1.** Patient characteristics in the pre-vaccination, vaccinated and unvaccinated cohorts.

| Characteristics |  | Pre-vaccination cohort<br>(1 Jan 2020 to 14 Dec 2021) |  | Vaccinated cohort<br>(1 June to 14 Dec 2021) |  | Unvaccinated cohort<br>(1 June to 14 Dec 2021) |  |
| --- | --- | --- | --- | --- | --- | --- | --- |
|  |  | N (%) | COVID-19 diagnoses | N (%) | COVID-19 diagnoses | N (%) | COVID-19 diagnoses |
| **All** |  | 15,211,471 | 827,074 | 11,822,640 | 750,370 | 2,851,183 | 147,044 |
| **Sex** | Female | 7,679,998 (50.5) | 451,321 | 6,233,098 (52.7) | 422,888 | 1,169,870 (41) | 78,520 |
|  | Male | 7,531,473 (49.5) | 375,753 | 5,589,542 (47.3) | 327,482 | 1,681,313 (59) | 68,524 |
| **Age, years** | 18-29 | 2,767,824 (18.2) | 216,367 | 1,570,150 (13.3) | 83,851 | 877,616 (30.8) | 39,918 |
|  | 30-39 | 2,819,706 (18.5) | 184,089 | 1,777,903 (15) | 135,148 | 875,004 (30.7) | 51,000 |
|  | 40-49 | 2,691,915 (17.7) | 168,773 | 1,988,754 (16.8) | 215,123 | 549,468 (19.3) | 33,780 |
|  | 50-59 | 2,766,992 (18.2) | 154,699 | 2,355,572 (19.9) | 179,657 | 321,157 (11.3) | 16,045 |
|  | 60-69 | 2,004,640 (13.2) | 66,193 | 1,876,023 (15.9) | 85,212 | 144,857 (5.1) | 4,629 |
|  | 70-79 | 1,496,715 (9.8) | 25,916 | 1,518,258 (12.8) | 39,372 | 58,785 (2.1) | 1,203 |
|  | 80-89 | 582,058 (3.8) | 8,685 | 625,214 (5.3) | 10,195 | 19,557 (0.7) | 396 |
|  | 90+ | 81,621 (0.5) | 2,352 | 110,766 (0.9) | 1,812 | 4,739 (0.2) | 73 |
| **Ethnicity** | White | 11,952,029 (78.6) | 621,492 | 9,749,555 (82.5) | 640,774 | 1,778,150 (62.4) | 112,297 |
|  | Mixed | 173,025 (1.1) | 11,646 | 106,632 (0.9) | 6,433 | 69,810 (2.4) | 3,657 |
|  | South Asian | 897,376 (5.9) | 91,523 | 601,551 (5.1) | 31,916 | 274,976 (9.6) | 9,069 |
|  | Black | 310,204 (2) | 21,313 | 172,108 (1.5) | 7,748 | 146,815 (5.1) | 7,064 |
|  | Other | 323,475 (2.1) | 16,378 | 196,904 (1.7) | 9,015 | 158,953 (5.6) | 3,854 |
|  | Missing | 1,555,362 (10.2) | 64,722 | 995,890 (8.4) | 54,484 | 422,479 (14.8) | 11,103 |
| **Index of multiple deprivation quintile** | 1: Most deprived | 2,881,855 (18.9) | 199,514 | 1,854,399 (15.7) | 112,517 | 851,173 (29.9) | 44,116 |
|  | 2 | 2,979,241 (19.6) | 177,895 | 2,179,460 (18.4) | 136,092 | 683,330 (24) | 34,762 |
|  | 3 | 3,278,622 (21.6) | 165,299 | 2,616,291 (22.1) | 161,666 | 570,594 (20) | 29,246 |
|  | 4 | 3,136,033 (20.6) | 151,449 | 2,617,451 (22.1) | 168,694 | 434,995 (15.3) | 22,492 |
|  | 5: Least deprived | 2,935,720 (19.3) | 132,917 | 2,555,039 (21.6) | 171,401 | 311,091 (10.9) | 16,428 |
| **Region** | East | 3,574,984 (23.5) | 190,225 | 2,803,629 (23.7) | 161,600 | 649,800 (22.8) | 33,968 |
|  | East Midlands | 2,684,304 (17.6) | 157,426 | 2,084,802 (17.6) | 138,836 | 470,137 (16.5) | 28,240 |
|  | London | 946,362 (6.2) | 52,269 | 596,361 (5) | 31,737 | 398,742 (14) | 10,583 |
|  | North East | 742,622 (4.9) | 50,348 | 568,359 (4.8) | 42,985 | 123,134 (4.3) | 7,159 |
|  | North West | 1,350,901 (8.9) | 89,301 | 1,060,641 (9) | 75,137 | 193,798 (6.8) | 11,782 |
|  | South East | 1,007,031 (6.6) | 40,924 | 821,312 (6.9) | 49,929 | 171,562 (6) | 8,780 |
|  | South West | 2,141,441 (14.1) | 63,860 | 1,841,113 (15.6) | 114,048 | 285,718 (10) | 17,460 |
|  |  | N (%) | COVID-19 diagnoses | N (%) | COVID-19 diagnoses | N (%) | COVID-19 diagnoses |
|  | West Midlands | 576,420 (3.8) | 42,691 | 399,516 (3.4) | 24,722 | 148,724 (5.2) | 7,417 |
|  | Yorkshire/Humber | 2,187,406 (14.4) | 140,030 | 1,646,907 (13.9) | 111,376 | 409,568 (14.4) | 21,655 |
| Smoking status | Never smoker | 7,105,315 (46.7) | 423,039 | 5,635,383 (47.7) | 372,057 | 1,174,015 (41.2) | 59,053 |
|  | Ever smoker | 4,798,071 (31.5) | 251,088 | 4,123,628 (34.9) | 280,924 | 560,038 (19.6) | 42,401 |
|  | Current smoker | 2,653,966 (17.4) | 115,929 | 1,671,805 (14.1) | 82,343 | 783,734 (27.5) | 37,074 |
|  | Missing | 654,119 (4.3) | 37,018 | 391,824 (3.3) | 15,046 | 333,396 (11.7) | 8,516 |
| Care home resident |  | 24,814 (0.2) | 3,854 | 28,229 (0.2) | 1,338 | 1,965 (0.1) | 69 |

There were 133,323, 32,466, and 2,140 incident T2DM diagnoses in the pre-vaccination, vaccination and unvaccinated cohorts, respectively (**Table 2**). Corresponding numbers of incident T1DM events were 14,917, 2410 and 45 respectively. In all cohorts, both T2DM and T1DM incidence was substantially greater after hospitalised COVID-19 than before or without COVID-19 or after non-hospitalised COVID-19. Among people in in the pre-vaccination cohort with follow-up data four months after diagnosis, there were 110,580 incident T2DM diagnoses of which 81,729 (74%) were defined as persistent T2DM. The proportion of T2DM diagnoses after COVID-19 defined as persistent was 3,186/4,385 (73%) and was similar after hospitalised COVID-19 (754/1,097 (69%)) and non-hospitalised COVID-19 (2,432/3,288 (74%)).

**Table 2.** Number of diabetes events in the pre-vaccination, vaccinated and unvaccinated cohorts, with person -years of follow-up, by COVID-19 severity. *Incidence rates are per 100,000 person-years.

| Event | COVID-19 severity | Pre-vaccination cohort (N=15,211,471) |  | Vaccinated cohort (N=11,822,640) |  | Unvaccinated cohort (N=2,851,183) |  |
| --- | --- | --- | --- | --- | --- | --- | --- |
|  |  | Event/person-years | Incidence rate* | Event/person-years | Incidence rate* | Event/person-years | Incidence rate* |
| Type 2 diabetes | No COVID-19 | 127,607/28,752,729 | 444 | 31,601/5,330,475 | 593 | 1,960/990,420 | 198 |
|  | Hospitalised COVID-19 | 1,247/29,582 | 4,215 | 134/1,474 | 9,092 | 116/1,264 | 9,174 |
|  | Non-hospitalised COVID-19 | 4,469/828,838 | 539 | 731/138,303 | 529 | 64/21,827 | 293 |
| Type 1 diabetes | No COVID-19 | 14,011/28,857,621 | 49 | 2,410/5,338,262 | 45 | 565/990,790 | 57 |
|  | Hospitalised COVID-19 | 54/30,993 | 174 | † | NA | 17/1,286 | 1,322 |
|  | Non-hospitalised COVID-19 | 852/833,322 | 102 | † | NA | 33/21,839 | 151 |
| Gestational diabetes | No COVID-19 | 18,742/14,362,707 | 130 | 1,414/2,831,550 | 50 | 1,451/390,451 | 372 |
|  | Hospitalised COVID-19 | 29/13,632 | 213 | † | NA | † | NA |
|  | Non-hospitalised COVID-19 | 1,013/454,467 | 223 | † | NA |  |  |
| Other or non-specified diabetes | No COVID-19 | 12,630/28,860,799 | 44 | 4,862/5,337,688 | 91 | 402/990,792 | 41 |
|  | Hospitalised COVID-19 | 70/31,000 | 226 | 7/1,507 | 464 | 6/1,287 | 466 |
|  | Non-hospitalised COVID-19 | 510/833,882 | 61 | 108/138,543 | 78 | 17/21,844 | 78 |
| **Restricted to participants with at least 4 months follow-up** |  |  |  |  |  |  |  |
| Type 2 diabetes | No COVID-19 | 106,195/28,752,729 | 369 |  |  |  |  |
|  | Hospitalised COVID-19 | 1,097/29,582 | 3,708 |  |  |  |  |
|  | Non-hospitalised COVID-19 | 3,288/828,838 | 397 |  |  |  |  |
| Persistent type 2 diabetes | No COVID-19 | 78,543/28,752,729 | 273 |  |  |  |  |
|  | Hospitalised COVID-19 | 754/29,582 | 2,549 |  |  |  |  |
|  | Non-hospitalised COVID-19 | 2,432/828,838 | 293 |  |  |  |  |

† insufficient events for estimation

### Incident T2DM after COVID-19 versus before or without COVID-19

In the pre-vaccination and unvaccinated cohorts, maximally adjusted HRs (aHRs) comparing T2DM incidence after COVID-19 with the incidence before or without COVID-19 were attenuated compared with age-, sex- and region-adjusted HRs (**Table 3**). However, maximally adjusted and age-sex adjusted aHRs were similar in the vaccinated cohort. In each cohort, T2DM incidence was elevated during the first four weeks after COVID-19 diagnosis: this elevation was greater in the unvaccinated cohort (aHR 4.82 [95% CI 3.65,6.36]) than the vaccinated cohort (1.42 [1.25,1.62]) (**Figure 1**). In the pre-vaccination cohort, T2DM incidence remained elevated 53-102 weeks post diagnosis (aHR 1.24 [1.12,1.37]). The elevation in T2DM incidence 5-28 weeks after COVID-19 was less marked in the vaccinated and unvaccinated cohorts (aHRs 1.17 [1.06,1.31] and 1.12 [0.74,1.68] respectively) than in the pre-vaccination cohort (1.30 [1.24-1.38]). In the pre-vaccination cohort, the aHRs for persistent T2DM were lower than for all T2DM during weeks 1-4, similar during weeks 5-28 and higher during weeks 29-52 (**Supplementary Figure 5**). In the unvaccinated cohort, aHRs for T2DM did not change markedly when censoring at first vaccination was removed (**Supplementary Table 5 and Supplementary Figure 6**).

**Figure 1.**
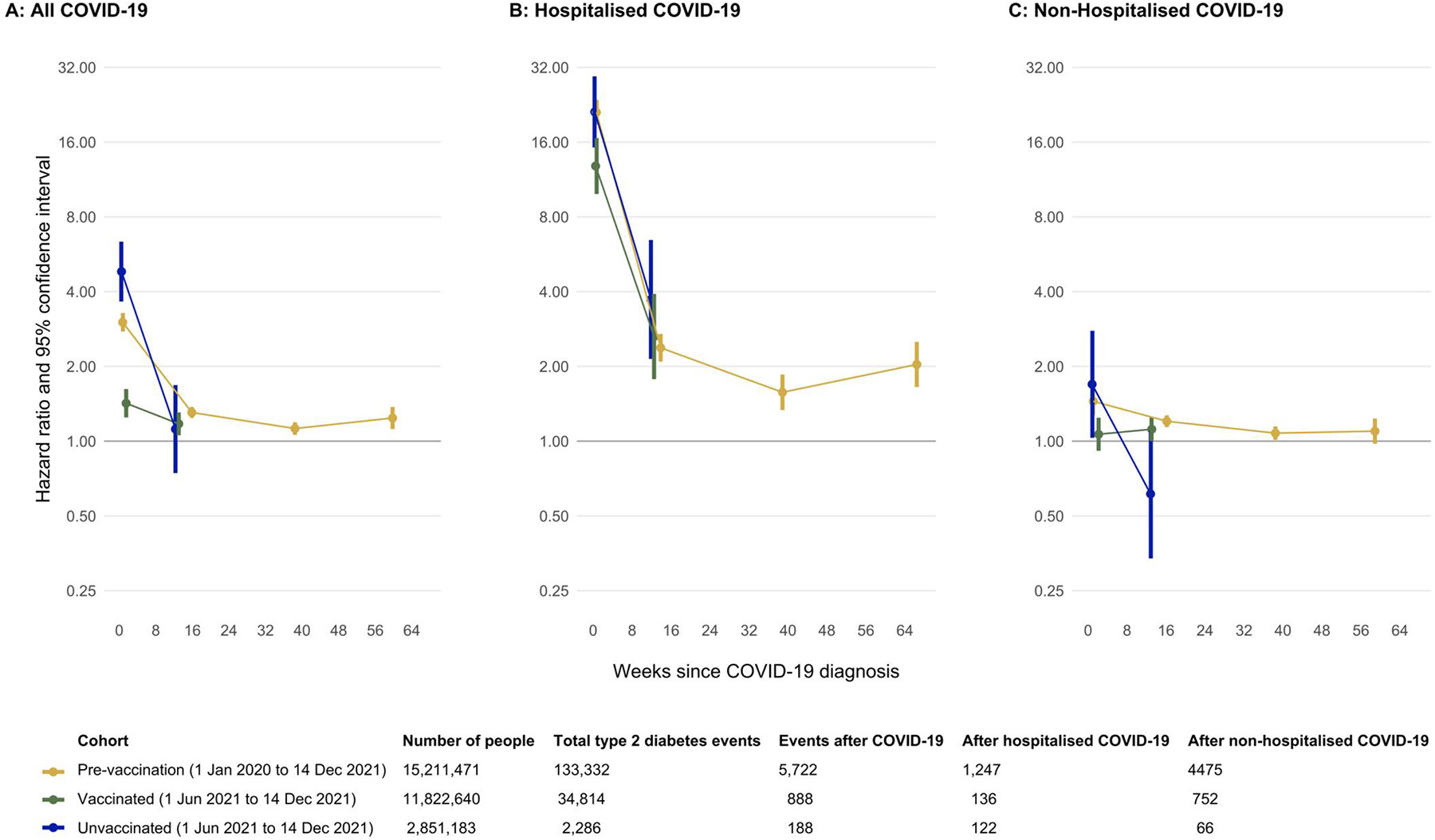
Maximally adjusted hazard ratios and 95% CIs comparing the incidence of type-2 diabetes events after COVID-19 (overall, hospitalised and non-hospitalised) with the incidence before or in the absence of COVID -19, in the pre-vaccination, vaccinated and unvaccinated cohorts. Points are plotted at the median time of the outcome event within each follow up period in each cohort.

**Table 3.**
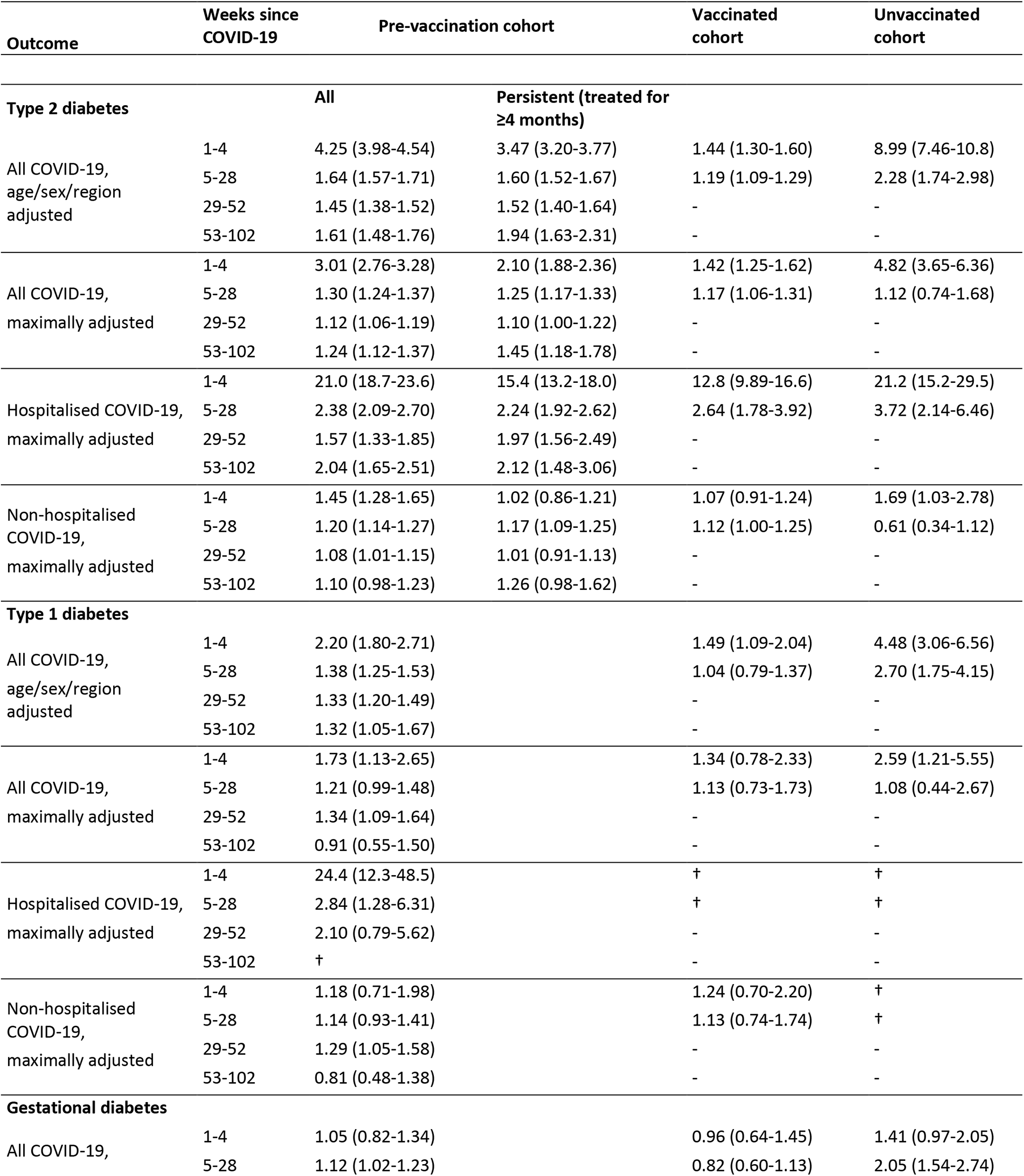
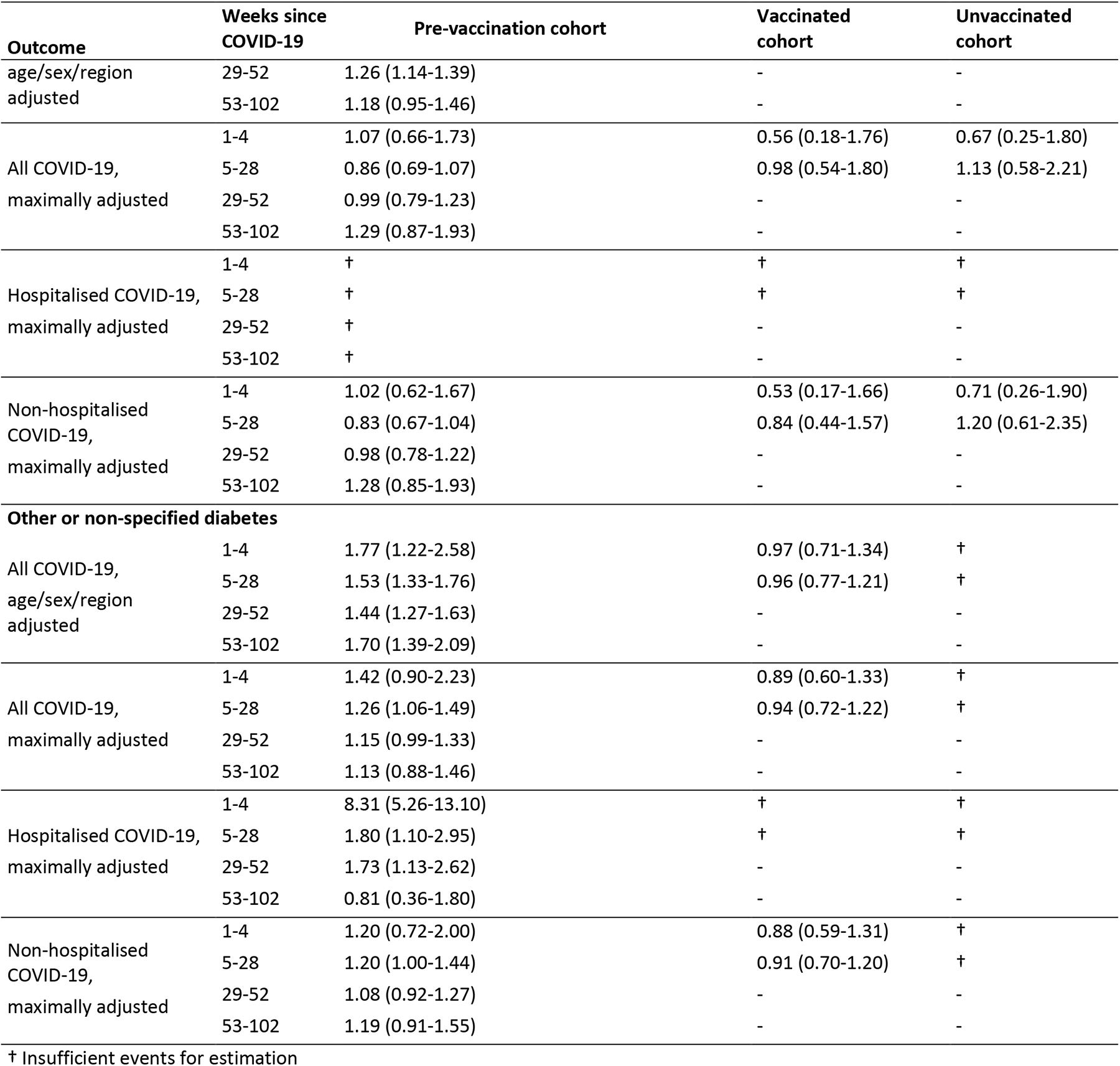
Adjusted hazard ratios (95% CI) comparing the incidence of diabetes events after COVID-19 with the incidence before or without COVID-19, in the pre-vaccination, vaccinated and unvaccinated cohorts, overall and according to COVID-19 severity. Hazard ratios are maximally adjusted unless otherwise stated.

The aHRs comparing T2DM incidence after COVID-19 with the incidence before or without COVID-19 were higher for hospitalised than non-hospitalised COVID-19, in each cohort (for example, 21.0 [18.7,23.6] versus 1.45 [1.28,1.65] in weeks 1-4 in the pre-vaccination cohort) (**Table 3**, **Figure 1**). T2DM incidence after hospitalised COVID-19 remained elevated beyond four weeks after COVID-19 diagnosis in each cohort. In the pre-vaccination cohort, the aHR for incident T2DM after hospitalised COVID-19 was 2.04 (1.65,2.51) during weeks 53-102. In the vaccinated cohort, T2DM incidence was not markedly elevated after non-hospitalised COVID-19. Patterns of aHRs estimated within shorter time intervals during the first 16 weeks after COVID-19 were similar to the main analyses (**Supplementary Figure 7**). The aHRs for persistent T2DM after hospitalised and non-hospitalised COVID-19 were consistent with those for all incident T2DM, except that the aHR for persistent T2DM after non-hospitalised COVID-19 was close to 1 (**Supplementary Figure 5**). In each cohort, and after both hospitalised and non-hospitalised COVID-19, a substantial proportion of T2DM diagnoses during weeks 1-4 were on the day of COVID-19 diagnosis (**Supplementary Table 6**) and the aHRs were markedly higher on the day of COVID-19 diagnosis than during the rest of weeks 1-4 (**Supplementary Figure 8)**.

The aHRs comparing T2DM incidence after COVID-19 with the incidence before or without COVID-19 were higher in older than younger age groups during weeks 1-4. Between age-group differences in aHRs were small in subsequent time periods (**Supplementary Table 7 and Supplementary Figure 9**). There were no marked differences between aHRS for T2DM by sex or between ethnic groups. In the pre-vaccination cohort, the aHR for T2DM during weeks 1-4 after COVID-19 diagnosis was higher for people without than with obesity, and for people without than with pre-diabetes. In both the vaccinated and unvaccinated cohorts, aHRs for T2DM in people with a COVID-19 diagnosis before the cohort start date could not be estimated because there were too few (<50) incident T2DM events after a further COVID-19 diagnosis during the cohort follow-up.

### Absolute excess risks of T2DM by age and sex

Estimated excess risks of T2DM 6 months post-COVID-19, standardised to the age and sex distribution of the pre-vaccination cohort, were 130, 56 and 94 per 100,000 people in the pre-vaccination, vaccinated and unvaccinated cohorts respectively (**Supplementary Figure 10**). Absolute differences in risk were higher in those aged over 60 years across, compared to younger age groups, in each cohorts. There was little difference in excess risk by sex, ethnicity, and presence or absence of pre-diabetes and obesity.

### Incident T1DM, gestational and other diabetes after COVID-19 versus before or without COVID-19

The aHRs comparing T1DM incidence after COVID-19 with the incidence before or without COVID-19 were higher during weeks 1-4 in the pre-vaccination and unvaccinated cohorts (1.73 [1.13,2.65] and 2.59 [1.21,5.55], respectively) than in the vaccinated cohort (1.34 [0.78-2.33]) (**Table 3 & Figure 2**)). In the pre-vaccination cohort, T1DM incidence remained elevated during weeks 29-52 (aHR 1.34 (1.09-1.64)) but not during weeks 53-102 (0.91 [0.55-1.50]). In the pre-vaccination cohort, aHRs for T1DM incidence were markedly higher after hospitalised COVID-19 than after non-hospitalised COVID-19. There was no consistent evidence of excess gestational diabetes incidence after COVID-19 in any group. The incidence of other types of diabetes was also elevated after COVID-19, most markedly during the first year after hospitalised COVID-19 in the pre-vaccination cohort (**Figure 2**).

**Figure 2.**
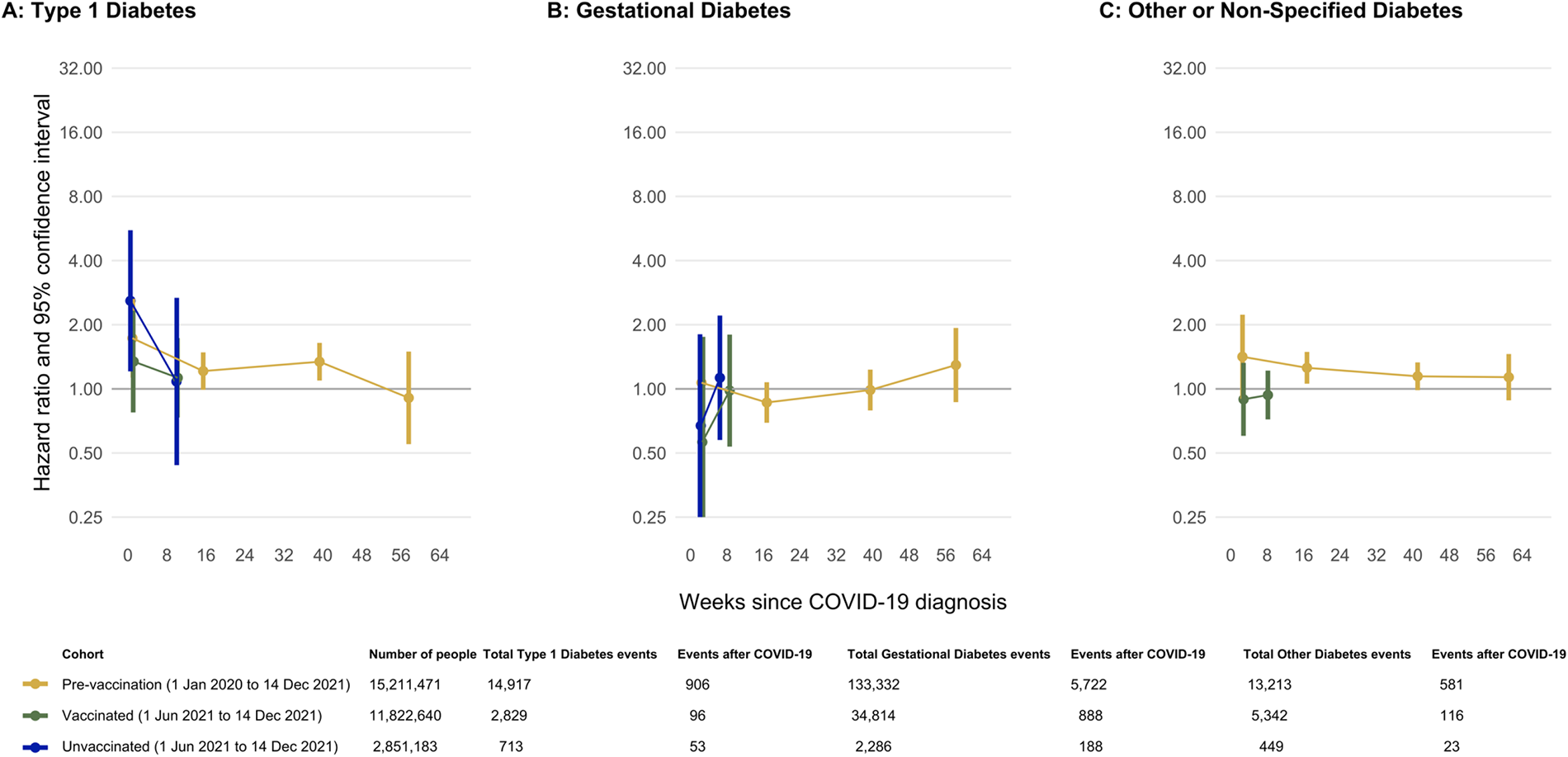
Maximally adjusted hazard ratios and 95% CIs comparing the incidence of diabetes events after COVID-19 (overall, hospitalised and non-hospitalised) with the incidence before or in the absence of COVID -19, in the pre-vaccination, vaccinated and unvaccinated cohorts. Points are plotted at the median time of the outcome event within each follow up period in each cohort.

## Discussion

### Key findings

In the cohort exposed before availability of COVID-19 vaccination, T2DM incidence was three times higher during the first four weeks after a diagnosis of COVID-19, compared with before or without COVID-19. T2DM incidence remained elevated by 24% overall during the second year after diagnosis, 2-fold higher in those hospitalised, and 10% higher in those not hospitalised with COVID-19. The majority of incident T2DM after COVID-19 was persistent. Elevation in T2DM incidence after COVID-19 was markedly attenuated in vaccinated compared with unvaccinated people (1.4-fold versus 4.8-fold during weeks 1-4 after COVID-19 diagnosis). T1DM incidence was only elevated during the first year after COVID-19 diagnosis. Gestational diabetes incidence did not appear elevated after COVID-19 diagnosis.

### Comparison with literature/ interpretation of findings

Elevated diabetes incidence (mostly T2DM) up to a year after COVID-19 has been reported in a recent systematic review^24^. However, the largest previous UK study did not confirm elevation in diabetes incidence beyond 12 weeks after COVID-19^6^. In that study, 20% of COVID-19 diagnoses were not laboratory confirmed, potentially attenuating the association between COVID-19 and incident diabetes. Additionally, people with pre-existing cardiovascular disease (CVD) were excluded, removing a group susceptible to both COVID-19 and T2DM.

There was little variation in post COVID-19 hazard ratios for incident T2DM between subgroups defined by ethnicity, sex, and pre-diabetes or obesity. This implies that absolute differences in excess risk after COVID-19 are higher in ethnic groups at higher risk of T2DM and in people with pre-diabetes and obesity.

Persistent T2DM, defined as being on glucose lowering medication or an HbA1c level consistent with T2DM at 4 months post diagnosis, was observed in 73% of incident T2DM after COVID-19, similar to the 74% persistence of incident T2DM before or without COVID-19. Using a similar definition of persistence, 56% of all newly diagnosed T2DM persisted up to a year post COVID-19 in a previous study ^25^. Additionally, 35% of newly diagnosed pre-diabetes post COVID-19 persisted at 6 months ^25^. These previous studies were in hospitalised patients: we found similar levels of persistence in those hospitalised and those not hospitalised with COVID-19.

No previous study has examined incident T2DM post COVID-19 diagnosis stratified by multiple periods of follow up; some excluded events in the first 30 days ^3^. Yet the very high T2DM incidence, including at the time of COVID-19 diagnosis in those hospitalised, should not be overlooked, given that the majority of these cases persist. Explanations for elevated T2DM incidence around the time of COVID-19 diagnosis include routine tests unmasking undiagnosed diabetes, or acute infection stress precipitating hyperglycaemia in those pre-disposed to diabetes. Incident diabetes later after COVID-19 diagnosis may reflect direct pancreatic beta-cell damage, or systemic inflammatory responses in those not previously at risk^26^. The hazard ratio for incident T2DM between 5-28 weeks in the vaccinated cohort (1.17) was lower than that for the pre-vaccination cohort (1.30) and almost identical to the hazard ratio of 1.16 observed for metabolic outcomes in a US cohort at 6 months^13^.

Pre-vaccination, T1DM incidence was elevated 1-4 weeks post COVID-19, and this elevation persisted during weeks 5-52, after both hospitalised and (to a lesser extent) non-hospitalised COVID-19, but not during the second year. This is consistent with previous studies in both adults (>18 years)^4^ and a meta-analysis of children and adolescents,^27^ but inconsistent with a Scottish study of children and young adults which only observed an excess risk in the month after COVID-19 ^11^. Thus, the balance of evidence does not support the hypothesis that elevated T1DM incidence soon after COVID-19 diagnosis is simply due to opportunistic testing for SARS-CoV-2 at the time of T1DM diagnosis or the short time lag between onset of T1DM and diagnosis ^15^. Previous studies largely censored follow up at around a year, and spanned the pre and post vaccination periods. We found that the elevation in T1DM incidence after COVID-19 was markedly attenuated in the vaccinated cohort.

We found no clear elevation in gestational diabetes incidence after COVID-19, though there was some indication of small increases in the incidence of other forms of diabetes which persisted beyond 9 months. We are not aware of any previous studies examining these sub-types.

### Strengths and limitations

A key strength is our sample size of around 15 million, enabling comparisons by diabetes type, vaccination status and time period post-infection in many population sub-groups, which no other study has done. Another strength is generalisability; the sample population was representative of the UK’s age, sex and ethnicity distribution. Linkage of primary and secondary care data with national SARS-CoV-2 testing data maximised and made more precise COVID-19 and other covariate ascertainment, and enabled study of hospitalised and non-hospitalised groups. All analyses accounted precisely for calendar time, so fluctuations in the incidence of COVID-19 and in the availability of health services will not have affected our findings.

Several limitations are present. Mild or asymptomatic COVID-19 cases will have been under-ascertained before widespread testing was available, biasing associations towards the null. Ascertainment of prevalent, rather than incident, diabetes may have been more likely in those with COVID-19, especially as diabetes pre-disposes to severe COVID-19. We addressed this possibility with sensitivity analyses excluding diabetes diagnoses on the same day as COVID-19 diagnosis. Further, elevated diabetes incidence after COVID-19 was evident in the non-hospitalised population, who were less likely to have diabetes tests in the acute period. Conversely, there may have been under ascertainment of diabetes due to reduced use of primary care services throughout the pandemic and cessation of normal screening activities, which would bias associations towards the null. Finally, misclassification of diabetes diagnoses is common in all settings ^21^ and may have particularly applied to acute hyperglycaemia in secondary care settings in this study. We used a previously published algorithm to overcome this ^21^.

### Clinical implications

Our findings have implications for the COVID-19, and potentially future, pandemics. England had around 2 million COVID-19 cases pre-vaccination, and 18 million post-vaccination ^28^. Taking account of non-vaccination, and using our estimates of absolute numbers of people with incident T2DM by vaccination status, we estimate around 14,000 additional new cases of T2DM 6 months post COVID-19 over the period of the pandemic. This contrasts with an estimated 56,000 new cases of all diabetes each year in England between 2015 and 2020 ^29^. Acknowledging the assumptions and uncertainties associated with such calculations, and that this is markedly lower than a previous estimate ^30^, this remains an alarmingly high number of new cases of T2DM, with significant costs to the individual and society. Encouraging vaccination, which markedly reduces risk of incident T2DM after COVID-19, is essential. Routine testing for diabetes after severe COVID-19, particularly in people at prior elevated risk of diabetes, and ensuring treatment and continued monitoring to identify those whose diabetes persists or resolves, should be considered. That COVID-19 appears to increase incidence of T1DM ^11^ and possibly T2DM ^31, 32^ to a greater extent than other respiratory infections adds weight to this recommendation. Our finding that T2DM incidence remains elevated up to 2 years after COVID-19 in the unvaccinated cohort emphasises the need to extend these analyses with longer follow up.

## Data Availability

All data were linked, stored, and analysed securely within the OpenSAFELY platform (https://opensafely.org/). Detailed pseudonymised patient data are potentially reidentifiable and therefore not shared. Details of access to OpenSAFELY secure data analytics platform is described on the OPENSAFELY (website https://opensafely.org/).

https://github.com/opensafely/post-covid-diabetes

## Contributors

### Funding

This study was supported by the COVID-19 Longitudinal Health and Wellbeing National Core Study, funded by the UKRI Medical Research Council (MC_PC_20059); the COVID-19 Data and Connectivity National Core Study, funded by the UKRI Medical Research Council; and by the CONVALESCENCE long COVID study, funded by the UK National Institute for Health and Care Research (COVID-LT-009). This work was also supported by Health Data Research UK, which is funded by the UK Medical Research Council, Engineering and Physical Sciences Research Council, Economic and Social Research Council, Department of Health and Social Care (England), Chief Scientist Office of the Scottish Government Health and Social Care Directorates, Health and Social Care Research and Development Division (Welsh Government), Public Health Agency (Northern Ireland), British Heart Foundation and Wellcome. SVE is funded by a Diabetes UK Sir George Alberti Research Training Fellowship (grant number 17/0005588). GC and AWo are supported by the British Heart Foundation (RG/13/13/30194; RG/18/13/33946), BHF Cambridge Centre of Research Excellence (RE/18/1/34212) and NIHR Cambridge Biomedical Research Centre (BRC-1215-20014; NIHR203312). RK and VW are supported by the Medical Research Council Integrative Epidemiology Unit at the University of Bristol [MC_UU_00011/1; MC_UU_00011/4]. RK, RD and JACS are supported by the NIHR Bristol Biomedical Research Centre and by Health Data Research UK South-West. YW was supported by an UKRI MRC Fellowship (MC/W021358/1) and received funding from UKRI EPSRC Impact Acceleration Account (EP/X525789/1). SI and AWo were funded by a British Heart Foundation–Turing Cardiovascular Data Science Award (BCDSA/100005). SI is funded by the International Alliance for Cancer Early Detection, a partnership among Cancer Research UK C18081/A31373, Canary Center at Stanford University, the University of Cambridge, OHSU Knight Cancer Institute, University College London, and the University of Manchester. RK and JM were supported by NIHR ARC West. RD and JACS were supported by Health Data Research UK. AWo is supported by the Stroke Association (SA CV 20/100018). AWo is part of the BigData@Heart Consortium, funded by the Innovative Medicines Initiative-2 Joint Undertaking under grant agreement No 116074. The views expressed are those of the authors and not necessarily those of the NIHR or the Department of Health and Social Care.

### Authors contributions

Author contributions are reported below in line with the Contributor Roles Taxonomy (CRediT).

Conceptualization: Nishi Chaturvedi, Rachel Denholm, Sophie Eastwood, Jonathan A C Sterne, Angela M Wood, Venexia Walker

Methodology: Nishi Chaturvedi, Rachel Denholm, Sophie Eastwood, Jonathan A C Sterne, Angela M Wood, Venexia Walker

Software: Louis Fisher, Jon Massey, Lisa Hopcroft, Tom Palmer, Simon Davy, Iain Dillingham, Catherine Murton, Felix Greaves, Ben Goldacre

Validation: Kurt Taylor, Venexia Walker, Genevieve Cezard, Rochelle Knight, Marwa Al Arab, Yinghui Wei, Elsie Horne, Lucy Teece, Jose Cuitun Coronado, Rachel Denholm

Investigation: Kurt Taylor, Venexia Walker, Genevieve Cezard, Rochelle Knight, Marwa Al Arab, Yinghui Wei, Elsie Horne, Lucy Teece, Harriet Forbes, Jose Cuitun Coronado, Sam Ip, Rachel Denholm

Data curation: Kurt Taylor, Venexia Walker, Genevieve Cezard, Rochelle Knight, Marwa Al Arab, Yinghui Wei, Elsie Horne, Lucy Teece, Harriet Forbes, Jose Cuitun Coronado, Rachel Denholm

Writing – Original Draft: Kurt Taylor, Sophie Eastwood, Nishi Chaturvedi, Jonathan A C Sterne, Rachel Denholm

Writing – Review & Editing: Kurt Taylor, Sophie Eastwood, Venexia Walker, Genevieve Cezard, Rochelle Knight, Marwa Al Arab, Yinghui Wei, Elsie Horne, Lucy Teece, Harriet Forbes, Alex Walker, Louis Fisher, Jon Massey, Lisa Hopcroft, Tom Palmer, Jose Cuitun Coronado, Sam Ip, Simon Davy, Iain Dillingham, Catherine Murton, Felix Greaves, John Macleod, Ben Goldacre, Angela Wood, Nishi Chaturvedi, Jonathan A C Sterne, Rachel Denholm

Visualization: Kurt Taylor, Venexia Walker, Genevieve Cezard, Rochelle Knight, Marwa Al Arab, Yinghui Wei, Elsie Horne, Lucy Teece, Jose Cuitun Coronado, Rachel Denholm

Project administration: Kurt Taylor, Jonathan A C Sterne, Venexia Walker, Rachel Denholm Funding acquisition: Nishi Chaturvedi, Angela M Wood, Jonathan A C Sterne

### Data sharing

Access to the underlying identifiable and potentially re-identifiable pseudonymised electronic health record data is tightly governed by various legislative and regulatory frameworks, and restricted by best practice. The data in OpenSAFELY is drawn from General Practice data across England where TPP is the data processor.

TPP developers initiate an automated process to create pseudonymised records in the core OpenSAFELY database, which are copies of key structured data tables in the identifiable records. These pseudonymised records are linked onto key external data resources that have also been pseudonymised via SHA-512 one-way hashing of NHS numbers using a shared salt. Bennett Institute for Applied Data

Science developers and PIs holding contracts with NHS England have access to the OpenSAFELY pseudonymised data tables as needed to develop the OpenSAFELY tools.

These tools in turn enable researchers with OpenSAFELY data access agreements to write and execute code for data management and data analysis without direct access to the underlying raw pseudonymised patient data, and to review the outputs of this code. All code for the full data management pipeline—from raw data to completed results for this analysis—and for the OpenSAFELY platform as a whole is available for review at https://github.com/OpenSAFELY.

## Acknowledgements

We are very grateful for all the support received from the TPP Technical Operations team throughout this work, and for generous assistance from the information governance and database teams at NHS England and the NHS England Transformation Directorate. We thank the CONVALESCENCE Study Long COVID PPIE group for their input and for sharing their experiences and expertise throughout the duration of the project.

## Information governance and ethical approval

NHS England is the data controller for OpenSAFELY-TPP; TPP is the data processor; all study authors using OpenSAFELY have the approval of NHS England. This implementation of OpenSAFELY is hosted within the TPP environment which is accredited to the ISO 27001 information security standard and is NHS IG Toolkit compliant (https://digital.nhs.uk/data-and-information/looking-after-information/data-security-and-information-governance/data-security-and-protection-toolkit).

Patient data are pseudonymised for analysis and linkage using industry standard cryptographic hashing techniques; all pseudonymised datasets transmitted for linkage onto OpenSAFELY are encrypted; access to the platform is via a virtual private network (VPN) connection, restricted to a small group of researchers; the researchers hold contracts with NHS England and only access the platform to initiate database queries and statistical models; all database activity is logged; only aggregate statistical outputs leave the platform environment following best practice for anonymisation of results such as statistical disclosure control for low cell counts (https://digital.nhs.uk/data-and-information/information-standards/information-standards-and-data-collections-including-extractions/publications-and-notifications/standards-and-collections/isb1523-anonymisation-standard-for-publishing-health-and-social-care-data).

The OpenSAFELY research platform adheres to the obligations of the UK General Data Protection Regulation (GDPR) and the Data Protection Act 2018. In March 2020, the Secretary of State for Health and Social Care used powers under the UK Health Service (Control of Patient Information) Regulations 2002 (COPI) to require organisations to process confidential patient information for the purposes of protecting public health, providing healthcare services to the public and monitoring and managing the COVID-19 outbreak and incidents of exposure; this sets aside the requirement for patient consent (https://web.archive.org/web/20200421171727/https://www.gov.uk/government/publications/covid-19-notification-to-gps-and-nhs-england-to-share-information). This was extended in November 2022 for the NHS England OpenSAFELY COVID-19 research platform (https://www.gov.uk/government/publications/coronavirus-covid-19-notification-to-organisations-to-share-information/coronavirus-covid-19-notice-under-regulation-34-of-the-health-service-control-of-patient-information-regulations-2002). In some cases of data sharing, the common law duty of confidence is met using, for example, patient consent or support from the Health Research Authority Confidentiality Advisory Group (https://www.hra.nhs.uk/about-us/committees-and-services/confidentiality-advisory-group/).

Taken together, these provide the legal bases to link patient datasets on the OpenSAFELY platform. GP practices, from which the primary care data are obtained, are required to share relevant health information to support the public health response to the pandemic, and have been informed of the OpenSAFELY analytics platform.

