## Supplemental material for "Diabetes following SARS-CoV-2 infection: Incidence, persistence, and implications of COVID-19 vaccination. A cohort study of fifteen million people"

**Tables and figures for supplementary material**

### Supplementary material

#### **Further details of statistical analyses**

If there were two events or fewer at any level of a potential confounder, the confounder was excluded from the analysis, after levels were aggregated when feasible. To make computations feasible, if the sample size was above four million, the datasets analysed included all individuals with the outcome event or who had COVID-19 during follow up, and a randomly selected subset of those without either the outcome or COVID-19 (“non-exposed controls”). The number of non-exposed controls sampled was based on the number of outcome events: 20 per event for less than 100,000 events, 10 per event for 100,000-500,000 events and 5 per event for >500,000 events. Analyses used inverse probability weights to account for this random sampling, and confidence intervals were derived using robust standard errors. All models were stratified by region, to account for between-region variation.

Absolute excess risks of T2D after COVID-19 were derived. The average daily incidence of each outcome before or without COVID-19 over the whole follow-up period was calculated, separately in subgroups defined by age group and sex. The incidence on each day after COVID-19 was derived by multiplying the daily incidence by the maximally adjusted HR for that day. Using a life table approach, age- and sex-specific cumulative risks over time, with and without COVID-19, were calculated, subtracting the latter from the former to get absolute excess risks over time after COVID-19 versus before or without COVID-19. The overall absolute excess risk was estimated using a weighted sum of the age- and sex-specific excess risks, weighted by the proportions of individuals in age and sex strata in the pre-vaccination cohort.

Individuals with missing age, sex, or deprivation are excluded from the analysis. We included a missing category for ethnicity. All other covariates are defined using the presence versus absence of specific codes, so have no identifiable missing values.

### **Supplementary Tables**

#### **Supplementary Table 1:** Derivation of diabetes outcomes codes in the OpenSAFELY environment. All code lists are available in the GitHub repo (<https://github.com/opensafely/post-covid-diabetes>).

| **Details** | **Link to primary care codelists** | **Link to secondary care codelists** |
| --- | --- | --- |
| Type 1 diabetes | <https://www.opencodelists.org/codelist/user/hjforbes/type-1-diabetes/674fbd7a/> | [OpenCodelists: Type 1 diabetes (secondary care)](https://www.opencodelists.org/codelist/opensafely/type-1-diabetes-secondary-care/2020-09-27/) |
| Type 2 Diabetes | <https://www.opencodelists.org/codelist/user/hjforbes/type-2-diabetes/3530d710/> | [OpenCodelists: Type 2 diabetes secondary care Bristol](https://www.opencodelists.org/codelist/user/r_denholm/type-2-diabetes-secondary-care-bristol/0b7f6cd4/) |
| Non-diagnostic | <https://www.opencodelists.org/codelist/user/hjforbes/nondiagnostic-diabetes-codes/50f30a3b/> |  |
| Other or non-specific | <https://www.opencodelists.org/codelist/user/hjforbes/other-or-nonspecific-diabetes/0311f0a6/> |  |
| Gestational diabetes | <https://www.opencodelists.org/codelist/user/hjforbes/gestational-diabetes/1ed423d1/> |  |

#### **Supplementary Table 2.** Derivation of confounder variables in the OpenSAFELY environment. All code lists are available in the GitHub repo (<https://github.com/opensafely/post-covid-diabetes>).

| **Confounder** | **Type** | **Definition** | **Data sources** |
| --- | --- | --- | --- |
| Sex | Categorical | Male, Female | Primary care |
| Age | Continuous | Modelled as age in years using a restricted cubic spline with 3 knots at the 10^th^, 50^th^ and 90^th^ percentiles | All |
| Ethnicity | Categorical | 1: White  2: Mixed  3: South Asian  4: Black  5: Other  6: Missing | All |
| Deprivation | Categorical | 10 categories from Index of Multiple Deprivation 2019 | Index of Multiple Deprivation |
| Region | Categorical | East of England  London  Midlands  Northeast and Yorkshire  Northwest  Southeast  Southwest | Primary care |
| Consultation rate | Continuous | Number of primary care contacts in the year prior to index date | Primary care |
| Smoking status | Categorial | E: Ever smoker  M: Missing  N: Never smoker  S: Current smoker | Primary care |
| Obesity | Binary | 1 if BMI>=30 or coded diagnosis for obesity; 0 otherwise | Primary care, HES APC |
| Acute myocardial infarction | Binary | 1 if diagnosis present; 0 otherwise | Primary care, HES APC |
| All stroke | Binary | 1 if diagnosis present; 0 otherwise | Primary care, HES APC |
| Other arterial embolism | Binary | 1 if diagnosis present; 0 otherwise | Primary care, HES APC |
| Venous thromboembolism events | Binary | 1 if diagnosis present; 0 otherwise | Primary care, HES APC |
| Heart failure | Binary | 1 if diagnosis present; 0 otherwise | Primary care, HES APC |
| Angina | Binary | 1 if diagnosis present; 0 otherwise | Primary care, HES APC |
| Dementia | Binary | 1 if diagnosis present; 0 otherwise | Primary care, HES APC |
| Liver disease | Binary | 1 if diagnosis present; 0 otherwise | Primary care, HES APC |
| Chronic kidney disease | Binary | 1 if diagnosis present; 0 otherwise | Primary care, HES APC |
| Cancer | Binary | 1 if diagnosis present; 0 otherwise | Primary care, HES APC |
| Hypertension | Binary | 1 if diagnosis or prescription present; 0 otherwise | Primary care, HES APC |
| Diabetes | Binary | 1 if diagnosis or prescription present; 0 otherwise | Primary care, HES APC |
| Depression | Binary | 1 if diagnosis present; 0 otherwise | Primary care, HES APC |
| Chronic obstructive pulmonary disease | Binary | 1 if diagnosis present; 0 otherwise | Primary care, HES APC |
| Healthcare worker | Binary | 1 if healthcare worker; 0 otherwise | NHS England COVID-19 data store (see: <https://docs.opensafely.org/study-def-variables/#cohortextractor.patients.with_healthcare_worker_flag_on_covid_vaccine_record>) |
| Care home resident | Binary | 1 if care home resident; 0 otherwise | Address matching CQC database (see: <https://docs.opensafely.org/study-def-variables/#cohortextractor.patients.care_home_status_as_of>)( |
| Total cholesterol/high-density lipoprotein [HDL] cholesterol ratio [TC/HDL] | Continuous | Data from the five year prior to start date will be used to derive TV/HDL ratio. TC/HDL values are derived from the recorded total and HDL cholesterol values.  Total Cholesterol values < 1.75 or > 20 and HDL values < 0.4 or > 5 were removed. | Primary care |
| BMI | Categorical | <18; 18-24; 25-29; 30+ | Primary care |
| History of prediabetes | Binary | Yes; no  Clinical diagnosis code | Primary care |
| History of gestational diabetes | Binary | Yes; no  Clinical diagnosis code | Primary care |

#### **Supplementary Table 3:** Summary of cohorts.

|  | **Pre-vaccination cohort** | **Vaccinated cohort** | **Unvaccinated cohort** |
| --- | --- | --- | --- |
| Start date | 01/01/2020, which is the approximate start date of the pandemic in the UK. | 01/06/2021, which is the date that the delta variant was thought to be ubiquitous in England. | 01/06/2021, which is the date that the delta variant was thought to be ubiquitous in England. |
| End date | 14/12/2021, which is the day that the UK Health Security Agency stated that over half of English cases they sampled have S Gene Target Failure, meaning they were likely Omicron, in [this report](https://assets.publishing.service.gov.uk/government/uploads/system/uploads/attachment_data/file/1042100/20211217_OS_Daily_Omicron_Overview.pdf). | 14/12/2021, which is the day that the UK Health Security Agency stated that over half of English cases they sampled have S Gene Target Failure, meaning they were likely Omicron, in [this report](https://assets.publishing.service.gov.uk/government/uploads/system/uploads/attachment_data/file/1042100/20211217_OS_Daily_Omicron_Overview.pdf). | 14/12/2021, which is the day that the UK Health Security Agency stated that over half of English cases they sampled have S Gene Target Failure, meaning they were likely Omicron, in [this report](https://assets.publishing.service.gov.uk/government/uploads/system/uploads/attachment_data/file/1042100/20211217_OS_Daily_Omicron_Overview.pdf). |
| Exclusion criteria | Patients will be excluded if they meet any of the following criteria:   - SARS-CoV-2 infection recorded prior to their index date | Patients will be excluded if they meet any of the following criteria:   - SARS-CoV-2 infection recorded prior to their index date (these individuals were required for a sensitivity analysis and so were not removed at the data extraction stage). - They do not have a record of two vaccination doses prior to the study end date. - They received a vaccination prior to 08-12-2020 (i.e., the start of the vaccination program). - They received a second dose vaccination before their first dose vaccination. - They received a second dose vaccination less than three weeks after their first dose. - They received mixed vaccine products before 07-05-2021 | Patients will be excluded if they meet any of the following criteria:   - SARS-CoV-2 infection recorded prior to their index date (these individuals were required for a sensitivity analysis and so were not removed at the data extraction stage). - They have a record of one or more vaccination doses prior to their index date - They could not be assigned to a vaccination group as defined by the Joint Committee on Vaccination and Immunisation (JCVI). |
| Follow-up start | Study start date. | Follow-up will start at the latest of the following dates (i.e., an individual’s index date):   - Two weeks after their second vaccination - Study start date | Follow-up will start at the latest of the following dates (i.e., an individual’s index date):   - 12 weeks after they became [eligible for vaccination](https://docs.google.com/spreadsheets/d/1Epre2Cv_4UVTwHJ6ccJN7QwDRq9pGGyQoWJoWolODZQ/edit?usp=sharing)) - Study start date |
| Follow-up end | Follow-up will end at the earliest of the following dates:   - Death - Outcome event - Study end date - Deregistration date | Follow-up will end at the earliest of the following dates:   - Death - Outcome event - Study end date - Deregistration date | Follow-up will end at the earliest of the following dates:   - Vaccination - Death - Outcome event - Study end date - Deregistration date |
| Cox regression time periods, full | [0,7), [7,14), [14,28), [28,56), [56,84), [84,197), [197, 365), [365,714) | [0,7), [7,14), [14,28), [28,56), [56,84), [84,197) | [0,7), [7,14), [14,28), [28,56), [56,84), [84,197) |
| Cox regression time periods, collapsed | [0,28), [28,197), [197, 365), [365,714) | [0,28), [28,197) | [0,28), [28,197) |

#### **Supplementary Table 4.** Patient medical characteristics in the pre-vaccination, vaccinated and unvaccinated cohorts.

| Medical characteristics | | Pre-vaccination cohort  (1 Jan 2020 to 14 Dec 2021) | | Vaccinated cohort (1 June to 14 Dec 2021) | | Unvaccinated cohort (1 June to 14 Dec 2021) | |
| --- | --- | --- | --- | --- | --- | --- | --- |
|  |  | **N (%)** | **COVID-19 diagnoses** | **N (%)** | **COVID-19 diagnoses** | **N (%)** | **COVID-19 diagnoses** |
| All |  | 15,211,471 | 827,074 | 11,822,640 | 750,370 | 2,851,183 | 147,044 |
| GP Consultations (in the past year) | 0 | 4,305,651 (28.3) | 189,147 | 2,977,723 (25.2) | 171,886 | 1,546,759 (54.2) | 45,578 |
|  | 1-6 | 5,941,109 (39.1) | 346,897 | 4,797,029 (40.6) | 320,304 | 800,844 (28.1) | 57,121 |
|  | 6+ | 4,964,711 (32.6) | 291,030 | 4,047,888 (34.2) | 258,180 | 503,580 (17.7) | 44,345 |
| Medical history | All stroke | 175,713 (1.2) | 6,221 | 184,464 (1.6) | 6,441 | 9,998 (0.4) | 544 |
|  | Acute myocardial infarction | 248,086 (1.6) | 8,534 | 252,478 (2.1) | 9,390 | 12,819 (0.4) | 678 |
|  | Angina | 384,099 (2.5) | 12,391 | 381,409 (3.2) | 13,785 | 15,318 (0.5) | 806 |
|  | Cancer | 4,587,644 (30.2) | 282,978 | 4,099,588 (34.7) | 322,046 | 540,802 (19) | 50,751 |
|  | Chronic kidney disease | 636,905 (4.2) | 21,083 | 654,537 (5.5) | 22,242 | 30,099 (1.1) | 1,773 |
|  | Chronic obstructive pulmonary disease | 373,426 (2.5) | 12,630 | 367,748 (3.1) | 13,552 | 20,708 (0.7) | 996 |
|  | Dementia | 79,398 (0.5) | 5,268 | 92,988 (0.8) | 2,863 | 2,966 (0.1) | 152 |
|  | Depression | 4,227,295 (27.8) | 239,894 | 3,507,144 (29.7) | 240,430 | 631,277 (22.1) | 47,017 |
|  | Gestational diabetes | 83,133 (0.5) | 6,436 | 75,970 (0.6) | 8,448 | 22,457 (0.8) | 2,555 |
|  | Healthcare worker | 492,814 (3.2) | 56,525 | 425,231 (3.6) | 34,475 | 17,819 (0.6) | 2,105 |
|  | Heart failure | 179,549 (1.2) | 6,277 | 203,638 (1.7) | 7,118 | 8,482 (0.3) | 471 |
|  | Hypertension | 4,366,805 (28.7) | 201,013 | 4,009,558 (33.9) | 212,657 | 371,505 (13) | 27,108 |
|  | Liver disease | 85,508 (0.6) | 3,937 | 78,230 (0.7) | 3,751 | 12,747 (0.4) | 650 |
|  | Other arterial embolism | 68,673 (0.5) | 2,677 | 83,727 (0.7) | 3,504 | 4,890 (0.2) | 282 |
|  | Venous thromboembolism | 216,643 (1.4) | 9,410 | 214,303 (1.8) | 11,140 | 21,482 (0.8) | 1,653 |

#### **Supplementary Table 5.** Adjusted hazard ratios (95% CI) comparing the incidence of type-2 diabetes events after COVID-19 with the incidence before or without COVID-19, in the unvaccinated cohort, overall and according to COVID-19 severity. Comparing results from the main analysis and to results where censoring at vaccination was removed.

| **Outcome** | **Weeks since COVID-19** | **Unvaccinated cohort** | **Unvaccinated cohort without censoring at vaccination** |
| --- | --- | --- | --- |
| All COVID-19, age/sex/region adjusted | 1-4 | 8.99 (7.46-10.8) | 7.78 (6.51-9.29) |
|  | 5-28 | 2.28 (1.74-2.98) | 2.25 (1.78-2.85) |
| All COVID-19, maximally adjusted | 1-4 | 4.82 (3.65-6.36) | 4.48 (3.44-5.83) |
|  | 5-28 | 1.12 (0.74-1.68) | 1.20 (0.85-1.70) |
| Hospitalised COVID-19, maximally adjusted | 1-4 | 21.2 (15.2-29.5) | 19.6 (14.2-27.1) |
|  | 5-28 | 3.72 (2.14-6.46) | 2.89 (1.67-5.01) |
| Non-hospitalised COVID-19, maximally adjusted | 1-4 | 1.69 (1.03-2.78) | 1.77 (1.13-2.75) |
|  | 5-28 | 0.61 (0.34-1.11) | 0.86 (0.55-1.34) |

#### **Supplementary Table 6.** Number of events and maximally adjusted hazard ratios (aHR), comparing the incidence of type 2 diabetes events after COVID-19 with the incidence before or without COVID-19, separately for day 0 (the day of COVID-19 diagnosis) and the rest of weeks 1-4 after COVID-19 diagnosis, in the pre-vaccination, vaccinated and unvaccinated cohorts, overall and according to COVID-19 severity.

|  | **Time since COVID-19** | **Pre-vaccination cohort** | | | **Vaccinated cohort** | | **Unvaccinated cohort** | |
| --- | --- | --- | --- | --- | --- | --- | --- | --- |
|  |  | **Number of events** | **aHR (95% CI)** | **Number of events** | | **aHR (95% CI)** | **Number of events** | **aHR (95% CI)** |
| All | Day 0 | 292 | 24.71 (21.22-28.77) | 63 | | 6.01 (4.4-8.2) | 45 | 29.21 (17.2-49.6) |
|  | Days 1-27 | 638 | 2.16 (1.95-2.39) | 281 | | 1.22 (1.06-1.41) | 79 | 3.73 (2.71-5.12) |
|  | Weeks 5-28 | 2411 | 1.30 (1.24-1.37) | 521 | | 1.17 (1.06-1.31) | 56 | 1.12 (0.74-1.68) |
|  | Weeks 29-52 | 1839 | 1.12 (1.06-1.19) | - | | - | - | - |
|  | Weeks 53-102 | 536 | 1.24 (1.12-1.37) | - | | - | - | - |
| Hospitalised COVID-19 | Day 0 | 159 | 147.95 (119.09-183.8) | 29 | | 96.49 (60.75-153.27) | 34 | 123.24 (63.98-237.42) |
|  | Days 1-27 | 380 | 15.85 (13.84-18.15) | 75 | | 9.29 (6.81-12.66) | 57 | 16.84 (11.58-24.49) |
|  | Weeks 5-28 | 373 | 2.38 (2.09-2.7) | 30 | | 2.64 (1.78-3.92) | 25 | 3.72 (2.14-6.45) |
|  | Weeks 29-52 | 217 | 1.57 (1.33-1.85) | - | | - | - | - |
|  | Weeks 53-102 | 118 | 2.04 (1.65-2.51) | - | | - | - | - |
| Non-hospitalised COVID-19 | Day 0 | 133 | 13.65 (11.04-16.88) | 34 | | 3.33 (2.19-5.07) | 11 | 12.21 (5.06-29.51) |
|  | Days 1-27 | 258 | 0.97 (0.83-1.14) | 206 | | 0.97 (0.82-1.14) | 22 | 1.22 (0.67-2.21) |
|  | Weeks 5-28 | 2038 | 1.20 (1.14-1.27) | 491 | | 1.12 (1-1.25) | 31 | 0.61 (0.34-1.11) |
|  | Weeks 29-52 | 1622 | 1.08 (1.01-1.15) | - | | - | - | - |
|  | Weeks 53-102 | 418 | 1.10 (0.98-1.23) | - | | - | - | - |

#### **Supplementary Table 7.** Maximally adjusted hazard ratios (95% CI) comparing the incidence of type-2 diabetes after COVID-19 with the incidence before or in the absence of COVID-19, in the pre-vaccination, vaccinated and unvaccinated cohorts, by subgroup.

| **Subgroup** | | **Weeks since COVID-19 diagnosis** | **Pre-vaccination cohort** | **Vaccinated cohort** | **Unvaccinated cohort** |
| --- | --- | --- | --- | --- | --- |
| Age group | 18-39 | 1-4 | 1.92 (1.34-2.76) | † | † |
|  |  | 5-28 | 1.19 (0.99-1.42) | † | † |
|  |  | 29-52 | 1.30 (1.07-1.59) | - | - |
|  |  | 53-102 | 1.28 (0.86-1.91) | - | - |
|  | 40-59 | 1-4 | 2.47 (2.19-2.78) | 1.08 (0.88-1.33) | 4.19 (2.90-6.07) |
|  |  | 5-28 | 1.30 (1.21-1.38) | 1.18 (1.02-1.37) | 1.20 (0.74-1.96) |
|  |  | 29-52 | 1.11 (1.03-1.20) | - | - |
|  |  | 53-102 | 1.07 (0.92-1.25) | - | - |
|  | 60-79 | 1-4 | 4.23 (3.69-4.84) | 1.63 (1.34-1.99) | † |
|  |  | 5-28 | 1.30 (1.18-1.43) | 1.19 (1.01-1.41) | † |
|  |  | 29-52 | 1.09 (0.98-1.21) | - | - |
|  |  | 53-102 | 1.53 (1.30-1.80) | - | - |
|  | 80-110 | 1-4 | 6.81 (4.8-9.67) | 5.05 (3.55-7.18) | † |
|  |  | 5-28 | 1.03 (0.74-1.43) | 1.34 (0.84-2.14) | † |
|  |  | 29-52 | 1.18 (0.88-1.6) | - | - |
|  |  | 53-102 | 1.13 (0.77-1.66) | - | - |
| Sex | Female | 1-4 | 2.57 (2.24-2.96) | 1.32 (1.08-1.62) | 4.49 (2.99-6.74) |
|  |  | 5-28 | 1.29 (1.2-1.39) | 1.15 (0.98-1.35) | 1.09 (0.61-1.95) |
|  |  | 29-52 | 1.15 (1.05-1.25) | - | - |
|  |  | 53-102 | 1.19 (1.03-1.38) | - | - |
|  | Male | 1-4 | 3.37 (3.02-3.76) | 1.48 (1.24-1.76) | 5.18 (3.55-7.55) |
|  |  | 5-28 | 1.32 (1.23-1.41) | 1.18 (1.03-1.36) | 1.15 (0.65-2.06) |
|  |  | 29-52 | 1.11 (1.02-1.2) | - | - |
|  |  | 53-102 | 1.28 (1.11-1.48) | - | - |
| Ethnicity | White | 1-4 | 3.01 (2.72-3.33) | 1.45 (1.25-1.66) | 4.55 (3.21-6.44) |
|  |  | 5-28 | 1.27 (1.19-1.35) | 1.18 (1.05-1.32) | 1.08 (0.65-1.82) |
|  |  | 29-52 | 1.12 (1.05-1.20) | - | - |
|  |  | 53-102 | 1.24 (1.10-1.40) | - | - |
|  | Black | 1-4 | 4.79 (3.36-6.84) | † | † |
|  |  | 5-28 | 1.63 (1.28-2.07) | † | † |
|  |  | 29-52 | 0.98 (0.69-1.38) | - | - |
|  |  | 53-102 | 1.50 (0.90-2.51) | - | - |
|  | South Asian | 1-4 | 2.84 (2.29-3.51) | 1.01 (0.58-1.74) | † |
|  |  | 5-28 | 1.35 (1.19-1.52) | 0.88 (0.57-1.34) | † |
|  |  | 29-52 | 1.11 (0.96-1.28) | - | - |
|  |  | 53-102 | 1.11 (0.87-1.42) | - | - |
|  | Other | 1-4 | 3.02 (1.60-5.67) | † | † |
|  |  | 5-28 | 1.27 (0.86-1.87) | † | † |
|  |  | 29-52 | 1.38 (0.91-2.08) | - | - |
|  |  | 53-102 | 1.63 (0.80-3.29) | - | - |
|  | Missing | 1-4 | 1.02 (0.42-2.46) | † | † |
|  |  | 5-28 | 1.66 (1.26-2.19) | † | † |
|  |  | 29-52 | 1.24 (0.88-1.75) | - | - |
|  |  | 53-102 | 1.19 (0.56-2.51) | - | - |
|  | Yes | 1-4 | 2.74 (2.46-3.05) | 1.37 (1.16-1.61) | 5.24 (3.83-7.18) |
| History of obesity |  | 5-28 | 1.21 (1.13-1.28) | 1.08 (0.94-1.23) | 0.96 (0.57-1.6) |
|  |  | 29-52 | 1.12 (1.04-1.2) | - | - |
|  |  | 53-102 | 1.28 (1.13-1.45) | - | - |
|  | No | 1-4 | 3.60 (3.13-4.15) | 1.47 (1.17-1.85) | 3.76 (2.11-6.72) |
|  |  | 5-28 | 1.50 (1.38-1.63) | 1.33 (1.12-1.59) | 1.52 (0.78-2.96) |
|  |  | 29-52 | 1.14 (1.03-1.26) | - | - |
|  |  | 53-102 | 1.18 (0.99-1.41) | - | - |
| History of pre-diabetes | Yes | 1-4 | 2.37 (2.01-2.79) | 1.18 (0.94-1.48) | † |
|  |  | 5-28 | 1.32 (1.21-1.45) | 1.34 (1.15-1.56) | † |
|  |  | 29-52 | 1.10 (0.99-1.22) | - | - |
|  |  | 53-102 | 1.29 (1.09-1.54) | - | - |
|  | No | 1-4 | 3.37 (3.05-3.73) | 1.57 (1.34-1.85) | 5.8 (4.25-7.93) |
|  |  | 5-28 | 1.3 (1.22-1.38) | 1.06 (0.92-1.23) | 1.23 (0.75-2.0) |
|  |  | 29-52 | 1.13 (1.06-1.22) | - | - |
|  |  | 53-102 | 1.22 (1.07-1.38) | - | - |
| † Insufficient events for estimation | | | |  |  |

### **Supplementary Figures**

#### **Supplementary Figure 1.** Diabetes type and presence adjudication algorithm.

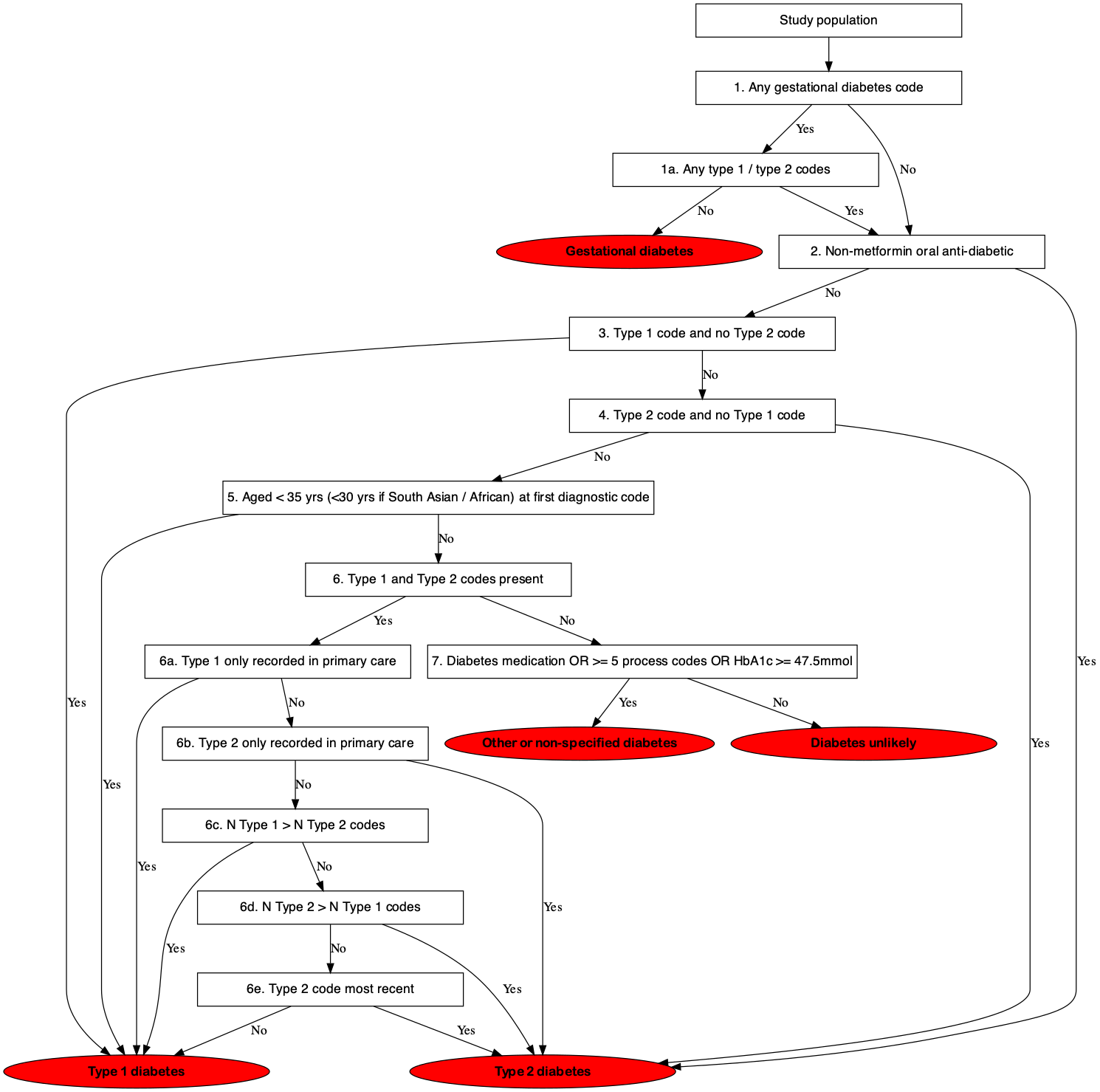

#### **Supplementary Figure 2.** Selection of study population in the pre-vaccination cohort.

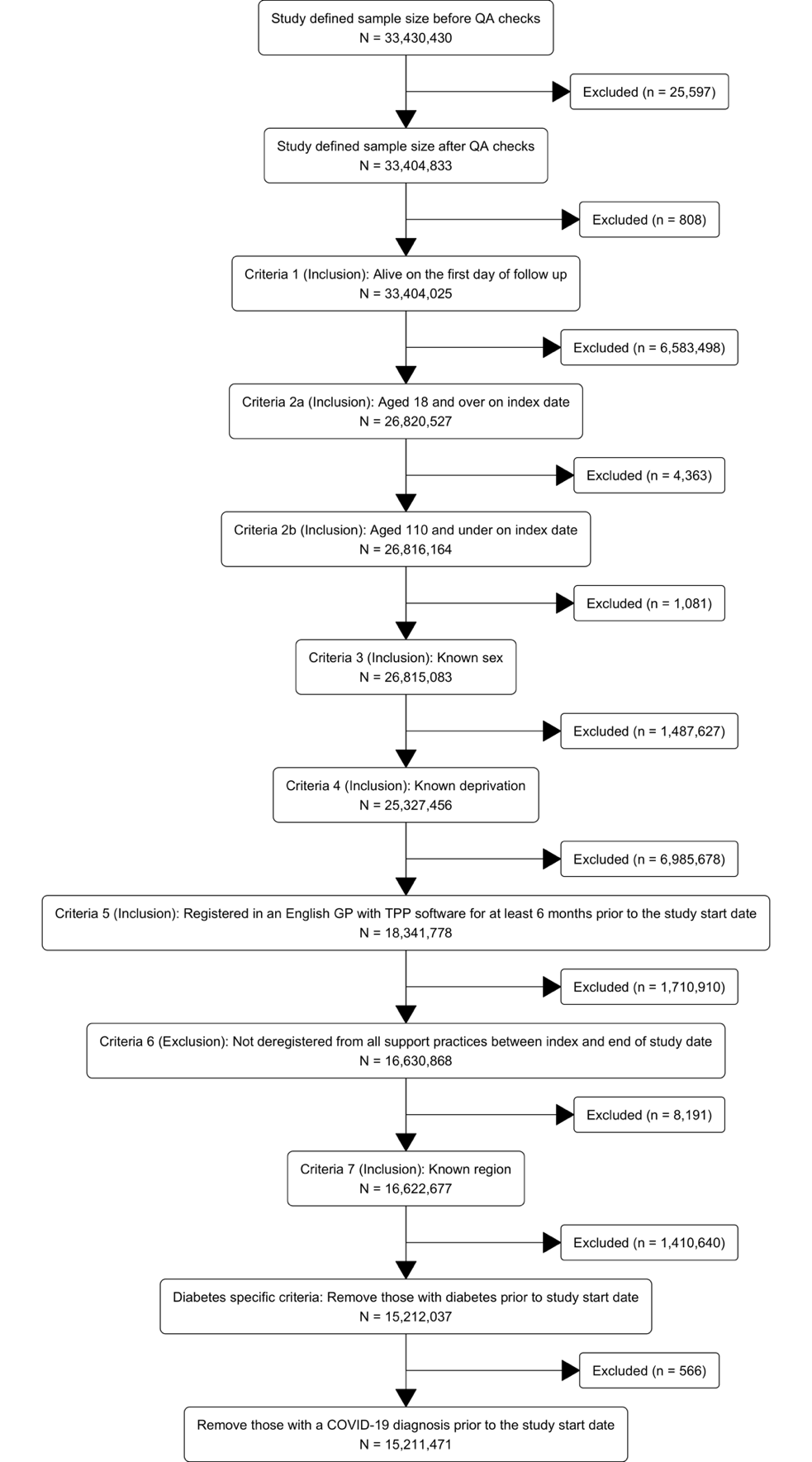

#### **Supplementary Figure 3.** Selection of study population in the vaccinated cohort.

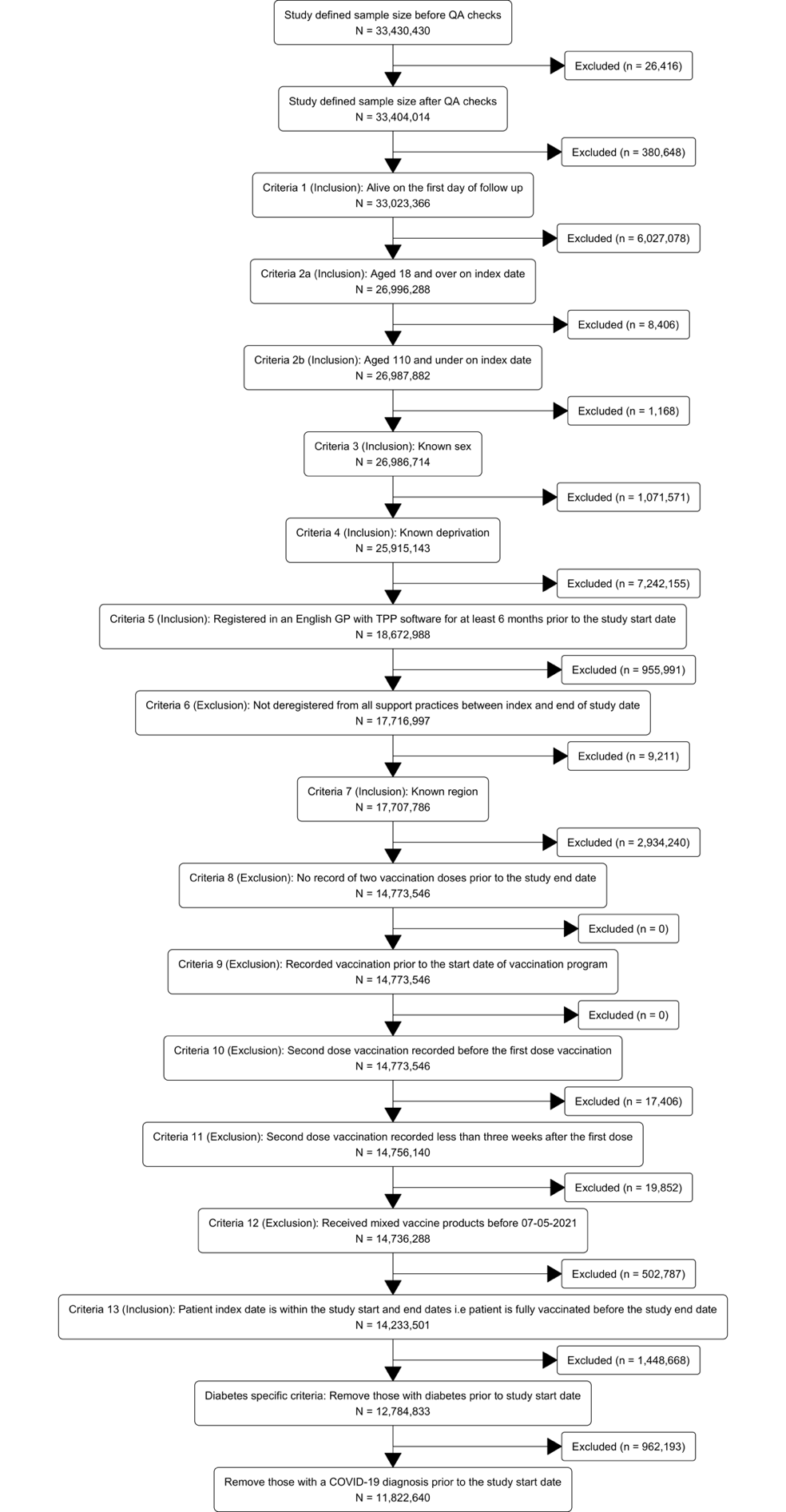

#### **Supplementary Figure 4.** Selection of study population in the unvaccinated cohort.

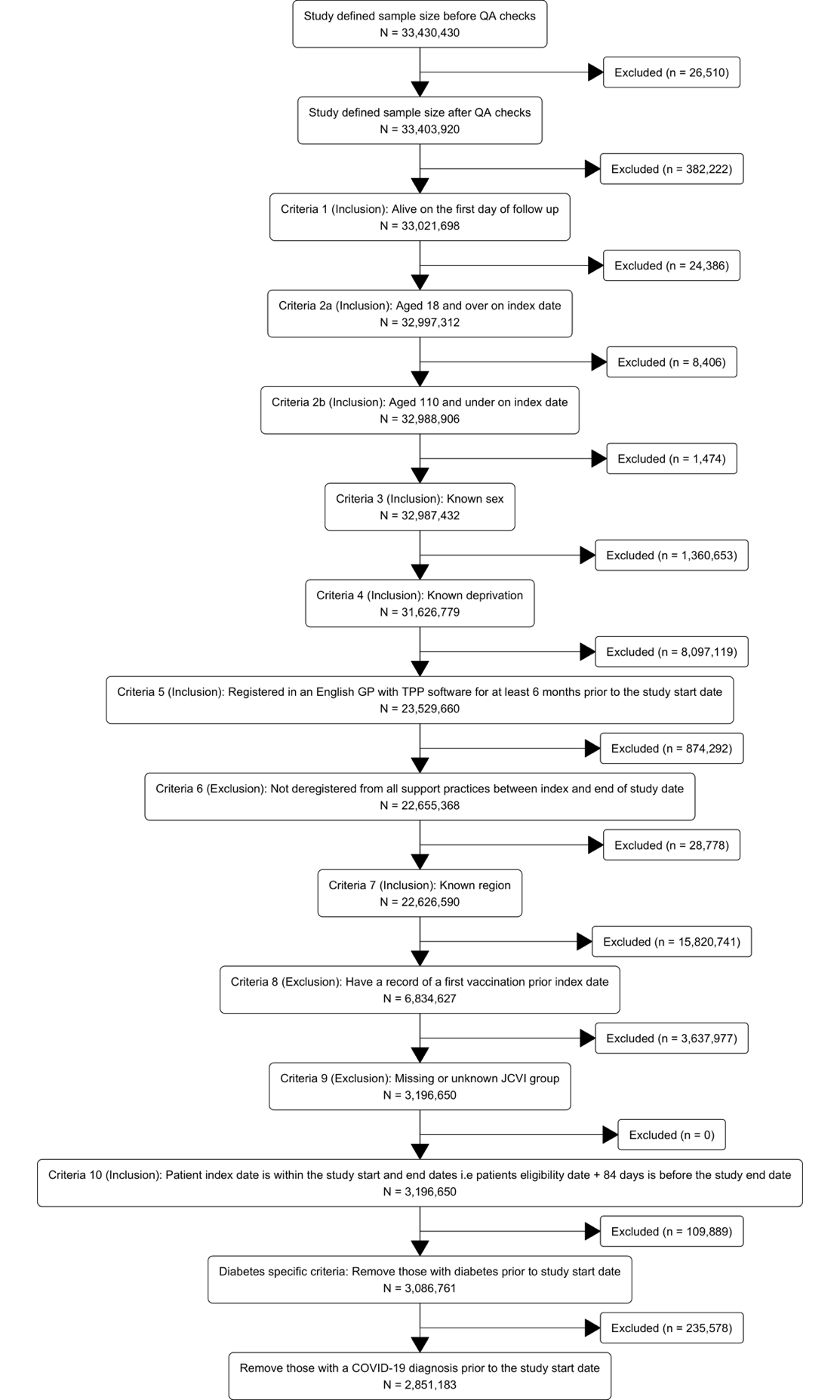

#### **Supplementary Figure 5.** Maximally adjusted hazard ratios and 95% CIs comparing the incidence of type-2 diabetes and persistent type-2 diabetes events after COVID-19 with the incidence before or without COVID-19, in the pre-vaccination cohort. Points are plotted at the median time of the outcome event within each follow up period comparing analyses for type-2 diabetes main analysis (as shown in Figure 1) and type-2 diabetes when restricted to cases that are still being treated after 4-months.

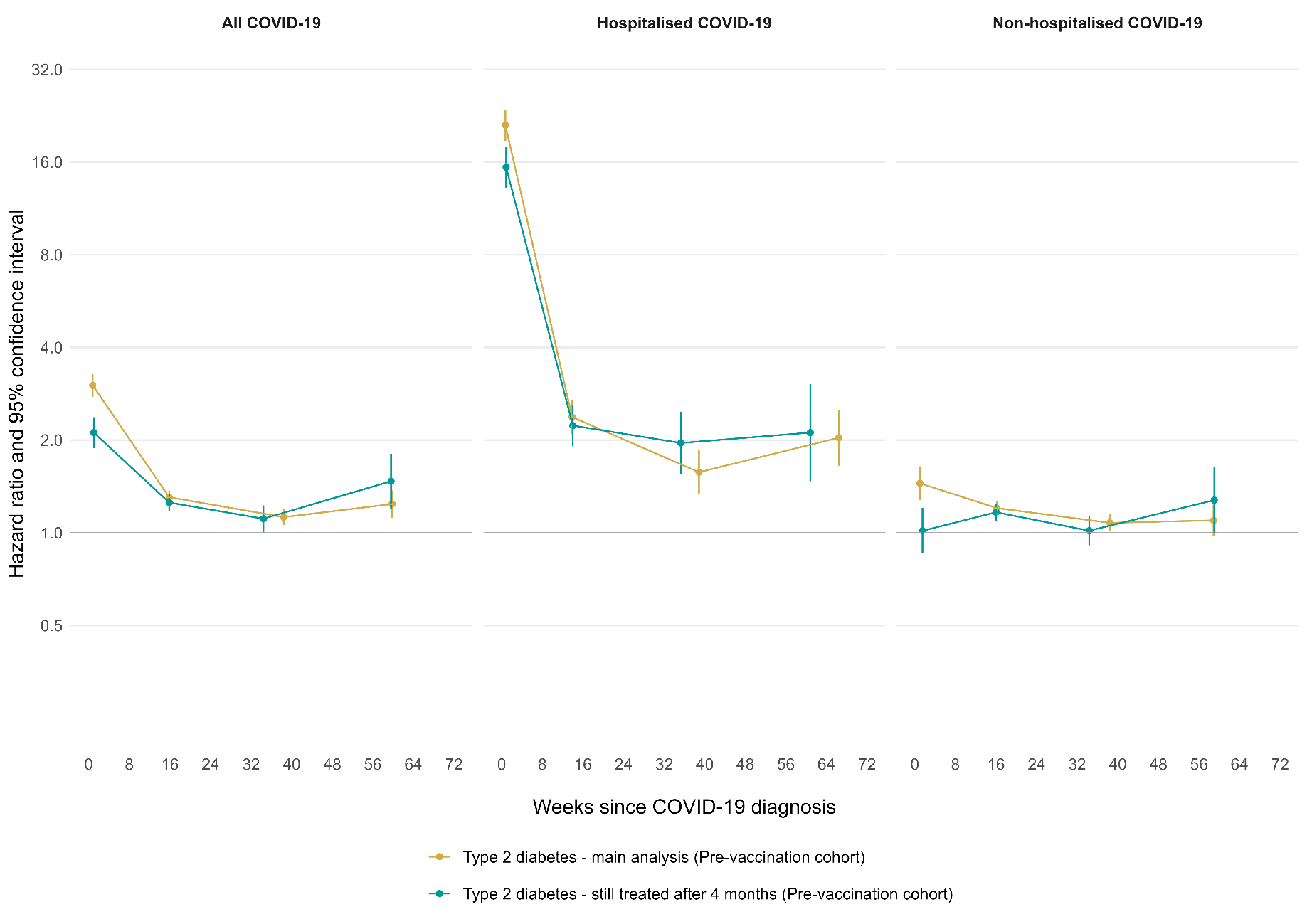
* N T2DM cases post-covid (%) with an end date >= 4 months from their t2dm diagnosis (end date calculated as minimum of death date, dereg date, cohort end date). ** N T2DM cases post-covid with 4 months follow up (%) that are “still being treated” (defined as a HBA1C >= 47.5 4 months from t2dm diagnosis OR has >= 2 prescriptions 4 months from t2dm diagnosis).

#### **Supplementary Figure 6.** Maximally adjusted hazard ratios and 95% CIs comparing the incidence of type-2 diabetes events after COVID-19 with the incidence before or without COVID-19, in the pre-vaccination cohort, comparing the main analysis to a sensitivity analysis censoring participants at vaccination eligibility.

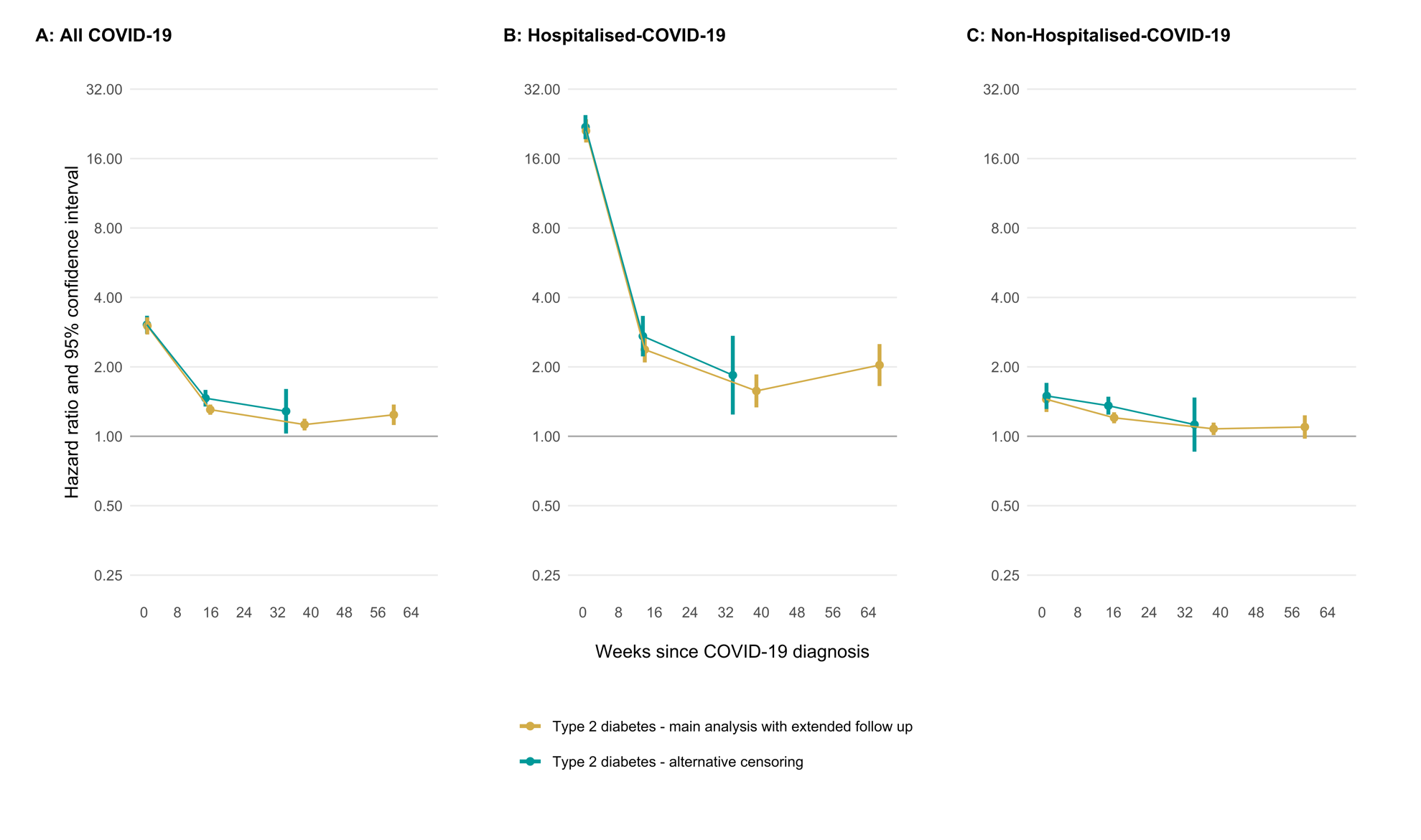

#### **Supplementary Figure 7.** Maximally adjusted hazard ratios and 95% CIs comparing the incidence of type-2 diabetes events after COVID-19 with the incidence before or without COVID-19, in the pre-vaccination and vaccinated cohorts, with shorter time intervals. Points are plotted at the median time of the outcome event within each follow up period in each cohort.

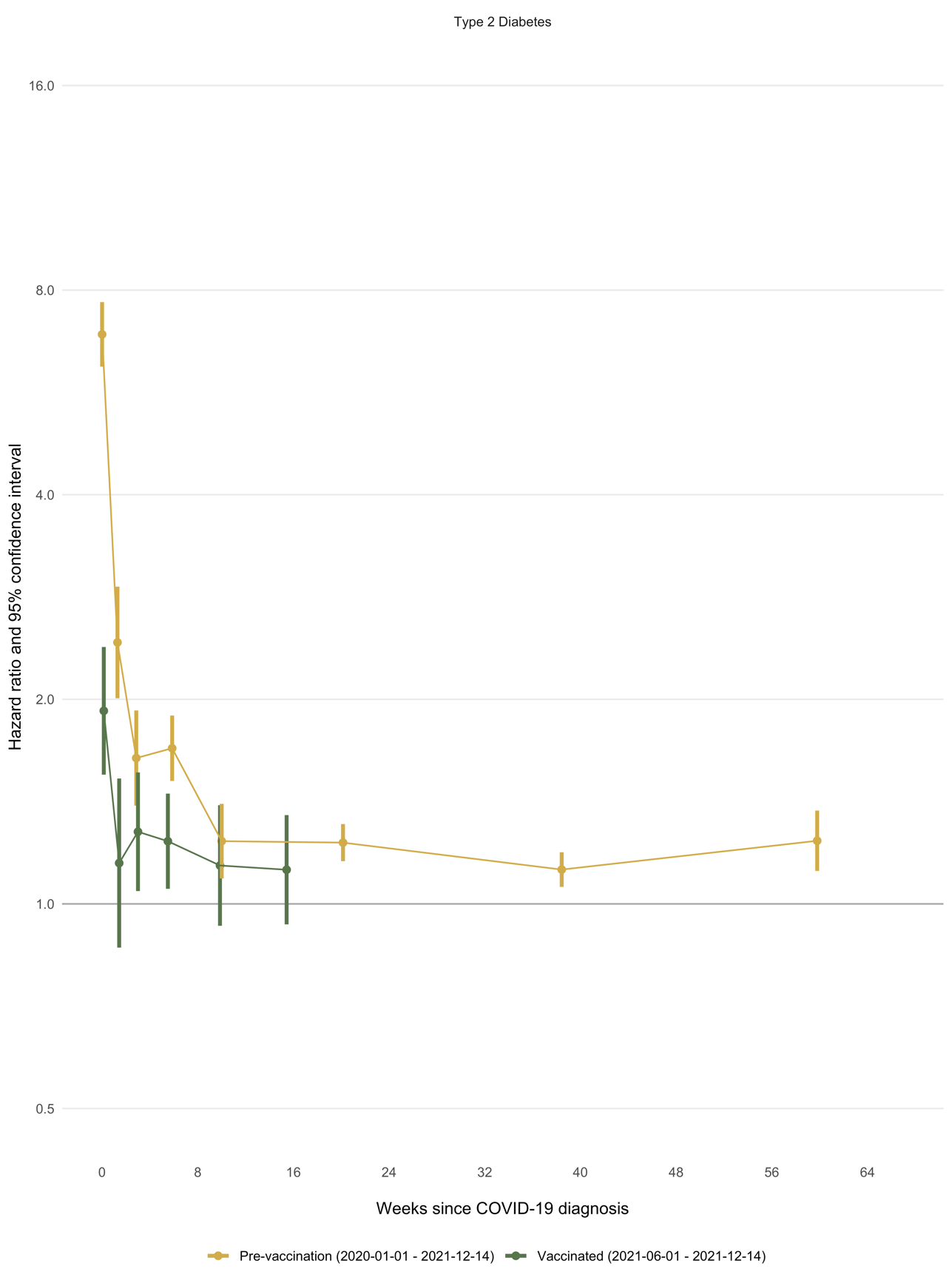

#### **Supplementary Figure 8.** Maximally adjusted hazard ratios comparing the incidence of type 2 diabetes events after COVID-19 with the incidence before or without COVID-19, separately for day 0 (the day of COVID-19 diagnosis) and the rest of weeks 1-4 after COVID-19 diagnosis, in the pre-vaccination, vaccinated and unvaccinated cohorts.

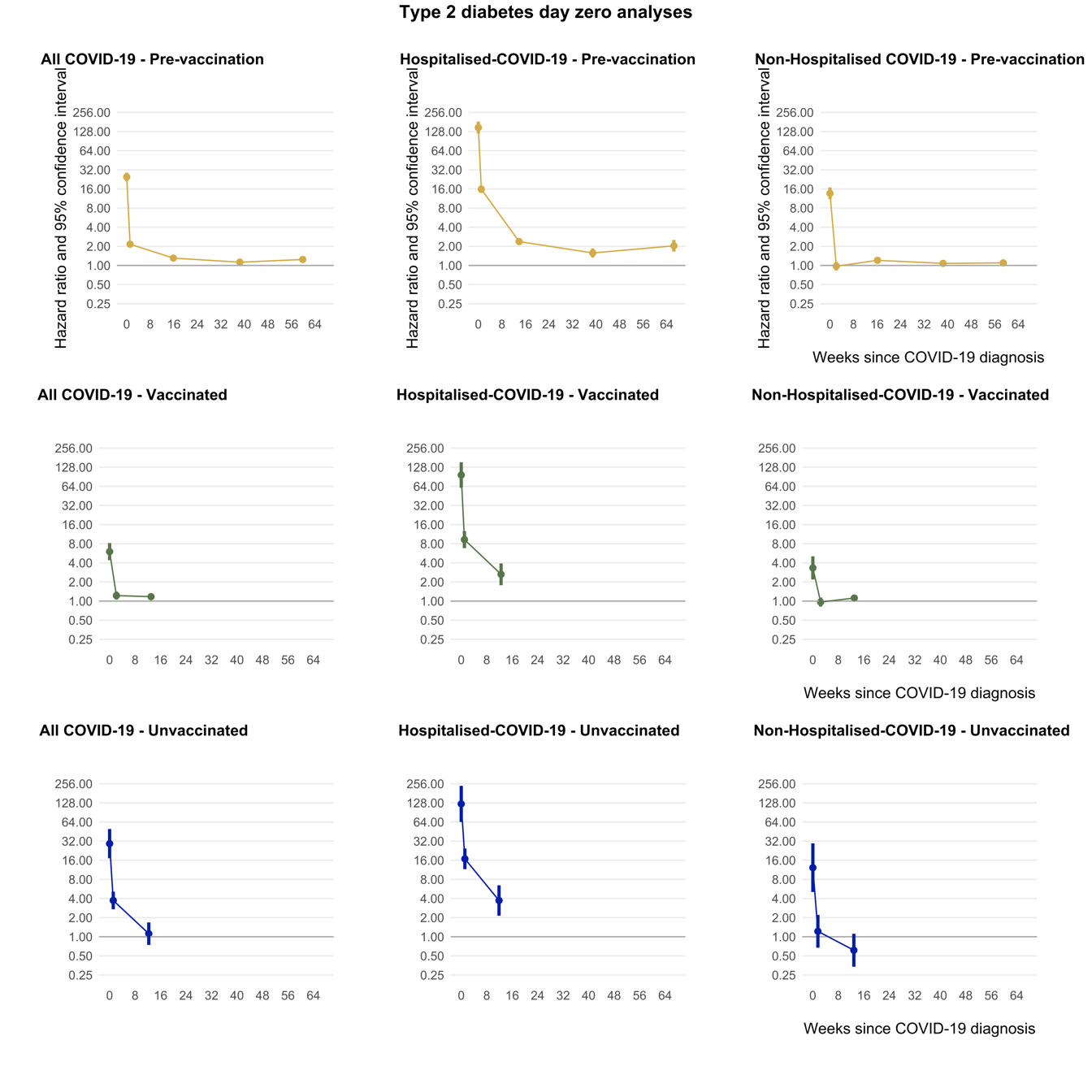

#### **Supplementary Figure 9.** Maximally adjusted hazard ratios and 95% CIs comparing the incidence of type-2 diabetes after COVID-19 with the incidence before or without COVID-19, in the pre-vaccination, vaccinated and unvaccinated cohorts, overall and by COVID-19 severity. Points are plotted at the median time of the outcome event within each follow up period in each cohort and are separated by subgroup (labelled on each individual plot)

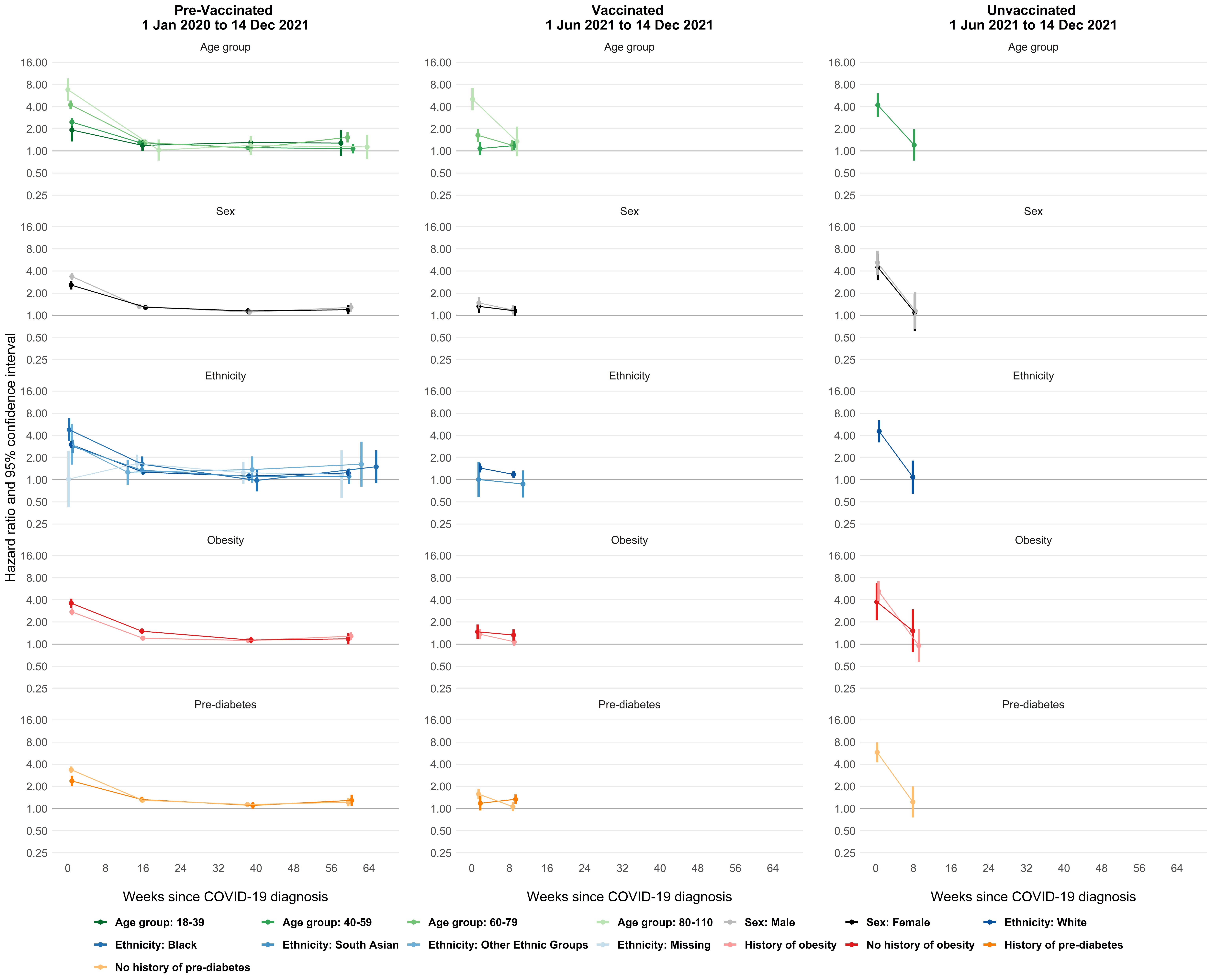

#### **Supplementary Figure 10.** Estimated absolute increase in risk of type-2 diabetes over time since diagnosis of COVID-19, compared with no COVID-19 diagnosis, in the pre-vaccination, vaccinated and unvaccinated cohorts. Increases in risks were estimated within age groups, and the estimated overall increase in risk is the average of these, weighted according to the proportions in each age group in the pre-vaccination cohort.

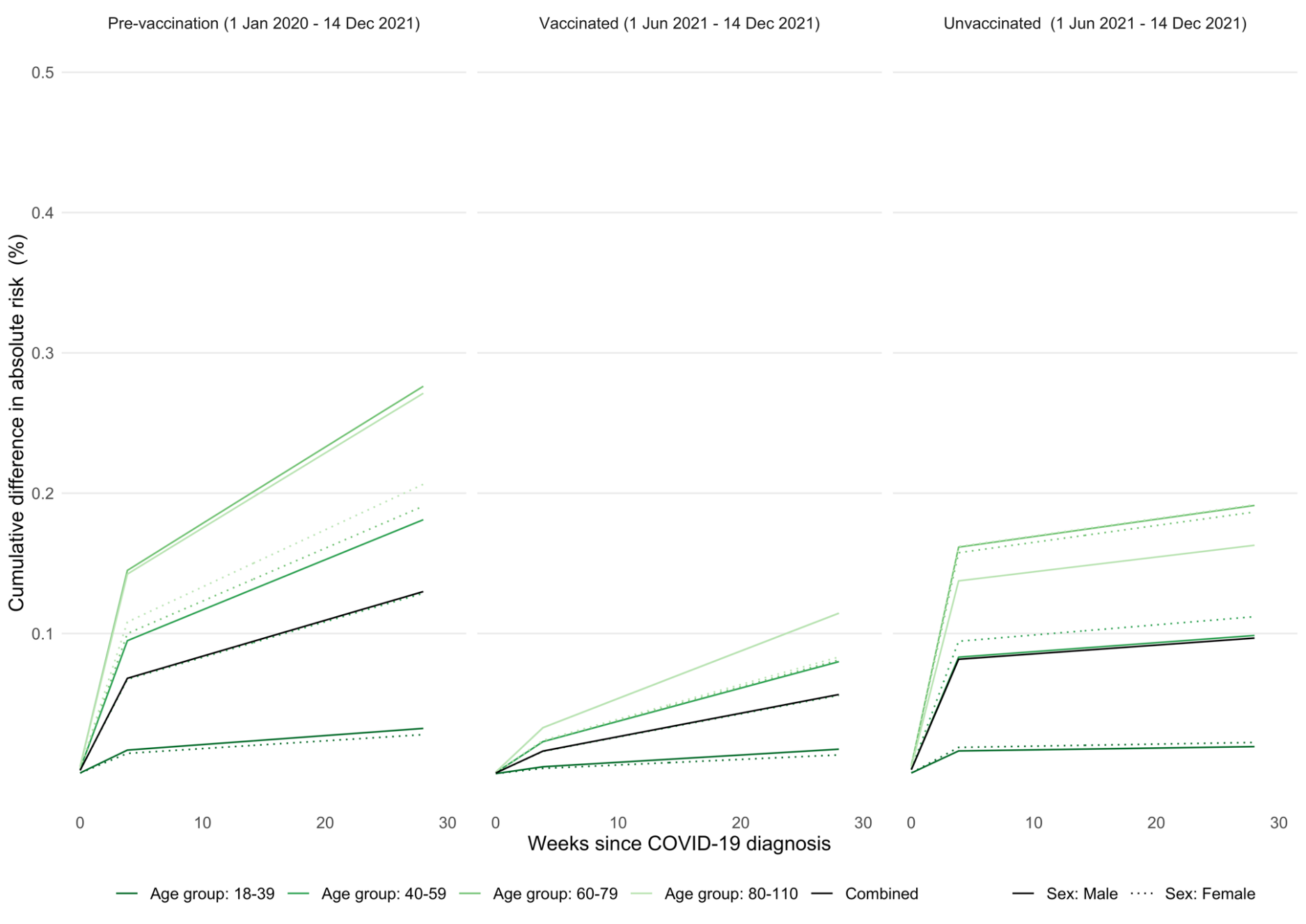
